# Heterogeneity of resting-state EEG features in juvenile myoclonic epilepsy and controls

**DOI:** 10.1101/2022.04.11.22273714

**Authors:** Amy Shakeshaft, Petroula Laiou, Eugenio Abela, Ioannis Stavropoulos, Mark P. Richardson, Deb K. Pal, the BIOJUME Consortium

**Affiliations:** Department of Basic & Clinical Neurosciences, Institute of Psychiatry, Psychology & Neuroscience, King’s College London, UK; MRC Centre for Neurodevelopmental Disorders, King’s College London, UK; Department of Biostatistics and Health Informatics, Institute of Psychiatry, Psychology & Neuroscience, King’s College London, UK; King’s College Hospital, London, UK; Evelina London Children’s Hospital, London, UK

**Keywords:** EEG, Epilepsy, Networks, Heterogeneity, Biomarkers

## Abstract

Abnormal EEG features are a hallmark of epilepsy, and abnormal frequency and network features are apparent in EEGs from people with idiopathic generalised epilepsy in both ictal and interictal states. Here, we characterise differences in the resting-state EEG of individuals with juvenile myoclonic epilepsy (JME) and assess factors influencing the heterogeneity of these EEG features. We collected EEG data from 147 participants with JME through the Biology of Juvenile Myoclonic Epilepsy (BIOJUME) study. 95 control EEGs were acquired from two independent studies (Chowdhury et al. (2014) and EU-AIMS Longitudinal European Autism Project). We extracted frequency and functional network-based features from 10-20s epochs of resting-state EEG, including relative power spectral density (PSD), peak alpha frequency, network topology measures and Brain Network Ictogenicity (BNI): a computational measure of the propensity of networks to generate seizure dynamics. The influence of covariates such as age, sex, antiseizure medication, EEG time and epoch length were investigated for each EEG feature prior to testing for differences between JME and control EEGs using univariate, multivariable and receiver operating curve (ROC) analysis. Additionally, associations of clinical phenotypes (seizure type, seizure control) with EEG features were investigated in the JME cohort. P-values were corrected for multiple comparisons. Univariate analysis showed significant differences in PSD in delta (2-5Hz) (p=0.0007, hedges’ g=0.55) and low-alpha (6-9Hz) (p=2.9×10^-8^, g=0.80) frequency bands, peak alpha frequency (p=0.000007, g=0.66), functional network mean degree (p=0.0006, g=0.48) and BNI (p=0.00006, g=0.56) between JME and controls. Since age (p=0.009) and epoch length (p=1.7×10^-8^) differed between the two groups and were potential confounders, we controlled for these covariates in multivariable analysis where disparities in EEG features between JME and controls remained. ROC analysis showed low-alpha PSD was optimal at distinguishing JME from controls, with an area under the curve of 0.72. Lower average normalized clustering coefficient and shorter average normalized path length were associated with poorer seizure control in JME patients. To conclude, individuals with JME have increased power of neural oscillatory activity at low-alpha frequencies, along with increased BNI compared to controls, supporting evidence from studies in other epilepsies with considerable external validity. In addition, the impact of confounders on different frequency-based and network-based EEG features observed in this study highlights the need for careful consideration and control of these factors in future EEG research in IGE particularly for their use as biomarkers.

## Introduction

An abnormal ictal EEG is the hallmark of epilepsy and the presence of epileptiform discharges, such as spontaneous or provoked 3-6Hz spike and waves, are routinely used to identify Idiopathic Generalised Epilepsy (IGE), including Juvenile Myoclonic Epilepsy (JME), in the clinical diagnostic process. A normal background EEG is required for a JME diagnosis,^1^ with no generalised or focal slowing.^2^ However, recent advances in computational analysis of EEG have shown that the interictal EEG of patients with IGE show differences compared to healthy controls.

Several studies on the EEG power spectrum in individuals with epilepsy have focused on alterations in the alpha rhythm: the oscillatory activity most apparent at posterior EEG electrodes at around 8-13Hz. Early EEG studies reported shifts in alpha power from a higher frequency (around 8-13Hz) to a lower frequency (6-8Hz) in patients with epilepsy,^3, 4^ with recent studies confirming these findings using robust and standardized quantitative methodology in patients with focal and generalised epilepsy, and controlling for antiseizure medication (ASM).^5–7^ Further, there is evidence for a decreased peak alpha frequency in patients with epilepsy^8^ and their asymptomatic first-degree relatives.^9^ The alpha rhythm is of particular interest in IGE due to evidence of its basis in cortico-thalamic interactions,^10^ which are central to the generation of generalised seizures/spike-wave discharges^11, 12^ and functionally and structurally altered in JME.^13, 14^ Aside from the alpha rhythm, there is evidence of abnormalities in other EEG frequency bands in IGE, such as increased theta^15, 16^ and beta power.^16^

Further, functional networks derived from EEG or MEG activity have altered topology in IGE, as quantified using graph theory,^17–21^ with evidence for the most extensive changes in functional connectivity existing in JME compared to other IGE syndromes.^22^ Whilst the reported differences in network topology are variable, increased clustering coefficient (indicating increased regularity of networks) is a somewhat consistent finding across studies.^20^ The variability of results in studies of functional connectivity in IGE likely comes from factors such as heterogeneity of patient groups, the age of participants, the frequency band in which the functional networks are derived, epoch length and the use of different network types (binary/weighted and undirected/directed), as well as methods used for their computation. Further, there is little known about the external validity and reproducibility of these network topology measures across different cohorts at different sites.

In addition to these static models of functional brain networks, dynamic models can help to further understand mechanisms of seizures by modelling transitions of brain networks from stable, interictal states to ictal states, and the parameters that are the most influential in this transition. Several dynamic models have been proposed as biomarkers of epilepsy, including the integration of global network structure and local node coupling into a phase oscillator model, which showed 57% sensitivity (given 100% specificity) and 66% specificity (given 100% sensitivity) as a biomarker of IGE.^19^ Brain Network Ictogenicity (BNI) is a computational method that quantifies the ability of a network to generate seizures. It models the dynamics at each node of a functional network using a dynamical model^23, 24^ and measures the time each network spends in a seizure-like state. Therefore, BNI depicts the propensity of a given functional network to generate seizure activity. This method has been used to predict the surgical outcome^24, 25^ and aid with resection site choice in patients with focal epilepsy,^26^ as well as differentiating between focal and generalised epilepsies^18^ when applied to EEG data. Further, using MEG data, Lopes et al. (2021)^27^ showed that BNI can act as a biomarker of JME with 73% classification accuracy. However, this model has not yet been explored in EEG data of patients with JME.

There have been variable findings as to whether these EEG features differ within IGE cohorts, according to specific phenotypes. Abela et al. (2019)^5^ showed that patients with poor seizure control (both IGE and focal epilepsy) had an increased shift of alpha oscillatory activity from high frequencies to low frequencies compared to those with good seizure control and healthy control participants. However, similar work by Pegg et al. (2020)^7^ found no difference in spectral power between patients with well-controlled IGE and drug resistant IGE. Additionally, there were no differences in functional network topology in the same IGE cohort.^21^ However, studies in patients in focal epilepsy have shown that dynamic network measures, such as BNI, show promise as a predictor of prognostic outcome,^24, 25^ as well as indicating differences in seizure/epilepsy type.^18^

The aim of this study is to assess and compare a range of resting-state interictal EEG features in a large cohort of individuals with JME to healthy controls, using EEGs collected across multiple sites. Further we will investigate the reliability and heterogeneity of these measures in these cohorts and any clinical factors influencing them. Table 1 shows the hypothesised direction of change of each EEG measure in JME compared to controls based on evidence from previous literature.

**Table 1.** Hypothesised change of direction of EEG features in JME compared to controls. BNI = Brain Network Ictogenicity; JME = Juvenile Myoclonic Epilepsy; PSD = Power spectral density.

| EEG measure | Hypothesised direction of change in JME compared to controls | Previous evidence |
| --- | --- | --- |
| Delta PSD | - | - |
| Low-alpha PSD | ↑ | Abela et al. (2019)<sup>5</sup><br>Pegg et al. (2020)<sup>7</sup> |
| High-alpha PSD | ↓ | Abela et al. (2019)<sup>5</sup> |
| Beta PSD | ↑ | Glaba et al. (2020)<sup>16</sup> |
| Peak alpha frequency | ↓ | Larsson & Kostov (2005)<sup>8</sup><br>Yaakub et al. (2020)<sup>9</sup> |
| Log<sub>10</sub> alpha shift | ↑ | Abela et al. (2019)<sup>5</sup> |
| Mean strength | ↑ | Chowdhury et al. (2014)<sup>17</sup> |
| Mean strength variance | ↑ | Chowdhury et al. (2014)<sup>17</sup> |
| Clustering coefficient | ↑ | Chowdhury et al. (2014)<sup>17</sup> |
| Path length | - | - |
| Small-world index | ↓ | Lee et al. (2020)<sup>22</sup> |
| BNI | ↑ | Lopes et al. (2021)<sup>27</sup> |

## Methods

### Participants

#### Biology of Juvenile Myoclonic Epilepsy study

The Biology of Juvenile Myoclonic Epilepsy (BIOJUME) Consortium is an international cross-sectional study, spanning 72 sites from 12 countries focused on young people and adults with JME. BIOJUME collects clinical and EEG data from patients with JME.

#### Participant recruitment

Inclusion criteria for this study are based on Avignon Class II consensus criteria for the diagnosis of JME^1^: (i) age of myoclonus onset 6-25 years; (ii) seizures comprising predominant or exclusive early morning myoclonus of upper extremities and (iii) EEG interictal generalised spikes/polyspike and waves with normal background. Participants aged between 6-55 years old were included. Exclusion criteria: (i) myoclonus only associated with carbamazepine or lamotrigine therapy; (ii) EEG showing predominant focal interictal epileptiform discharges or abnormal background; (iii) any evidence of progressive or symptomatic myoclonus epilepsy or focal seizures; (iv) global learning disability; (v) dysmorphic features or (vi) unable to provide informed consent.

### Clinical data collection

Clinical data were collected by study site research staff face-to-face and augmented by clinical records and EEG reports. The dataset included general demographics and health information, epilepsy history, including seizure types, seizure frequency, drug/lifestyle interventions and the presence of a photoparoxysmal response (PPR)^28^. Sites uploaded clinical data onto a secure central REDCap (Research Electronic Data Capture) database.^29, 30^

#### Phenotyping

A phenotyping panel, comprising seven epilepsy experts, then evaluated the diagnosis of JME according to the inclusion criteria, through consensus where necessary.

#### Seizure Prognosis

To test associations of EEG features with seizure prognosis, we categorised participants based on their answers to two questions: (i) whether they had been free from seizures over the past year and (ii) current ASM therapy, categorised as either no drug therapy, monotherapy (not necessarily the first appropriate ASM), dual therapy, or drug-resistant (two or more ASM failures). Based on answers to these two questions, participants were categorised based on their seizure control:

(a) **Good prognosis** - those who are seizure-free on monotherapy or no drug therapy,
(b) **Moderate prognosis** - those who are either seizure-free on ≥2 ASMs or not seizure-free on 0/1 ASM, or
(c) **Poor prognosis** - those who are not seizure-free on ≥2 ASMs (drug-resistant).

Using this classification, 8 individuals were unable to be categorised, due to missing data.

### EEG data collection

#### JME

Routine clinical EEGs were collected for participants from each BIOJUME study site. Natus Xltek or Nicolet systems were used for clinical EEG data collection. Sites used between 19-25 scalp electrodes placed according to the 10-20 system, apart from Danish EEGs which used the modified 10-20 system whereby electrodes T3/T4 are replaced by T7/T8 and electrodes T5/T6 are replaced by P7/P8. EEGs were sent securely to a central site where one (or two where possible) 10-20s resting-state, eyes-closed, awake segments with clear background oscillatory activity were selected by a trained EEG neurophysiologist.

#### Control

EEG control data were acquired from Chowdhury et al (2014)^17^ study and the EU-AIMS Longitudinal European Autism Project (LEAP) study.^31^

#### Chowdhury controls

Healthy control participants with no personal or family history of neurological or psychiatric diseases were recruited via a local research participant database. Participants were excluded if they had any other neuropsychiatric condition or a full-scale IQ <70. Ethical approval was obtained from King’s College Hospital Research Ethics Committee (08/H0808/157). Written informed consent was obtained from all participants. EEGs were acquired as previously described in Chowdhury et al (2014).^17^ Briefly, conventional 10-20, 19 channel scalp EEG were collected (sampling rate 256Hz, band-pass filtered 0.3-70Hz) using a NicoletOne system. A single 20s epoch was selected which included continuous dominant background rhythm with eyes closed, without any artefacts or patterns indicating drowsiness or arousal.

#### LEAP controls

Another control group was recruited as part of the EU-AIMS Longitudinal European Autism Project (LEAP).^31^ Only healthy control EEG data were used. Participants were typically developing individuals aged 6-30 years. 2 minutes of eyes closed resting-state EEG were recorded per participant. Data used for this study was acquired from three sites: King’s College London (KCL, United Kingdom) (N=2), University Campus BioMedico (UCBM, Rome, Italy) (N=15) and University Medical Centre Utrecht (UMCU, Netherlands) (N=40). The following EEG systems were employed: Brainvision (KCL), Biosemi (UMCU) and Micromed (UCBM), with sampling frequencies of 5000Hz (KCL), 2048Hz (UMCU) and 256-1000Hz (UCBM). All sites used 10-20 layout caps, with 60-64 electrodes. Eyes closed, resting-state segments were marked on the EEGs and from these segments, 10-20 second resting-state, eyes-closed, awake segments with clear background oscillatory activity were selected.

### EEG pre-processing

An overview of the EEG processing and analysis pipeline can be seen in Figure 1.

**Figure 1.**
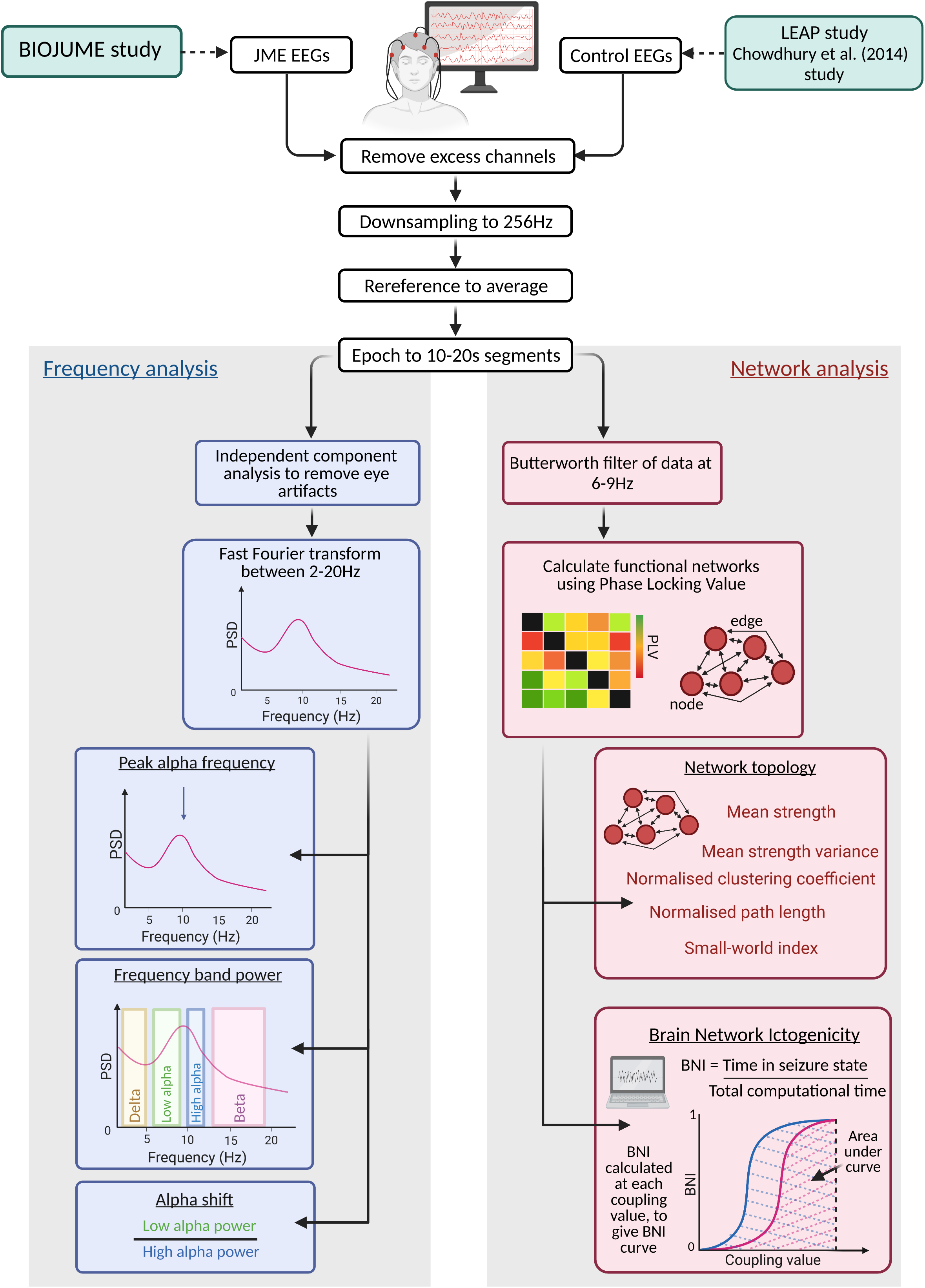
Summary of methods for EEG processing and analysis. BIOJUME = Biology of Juvenile Myoclonic Epilepsy; BNI = Brain Network Ictogenicity; JME = Juvenile myoclonic epilepsy; LEAP = Longitudinal European Autism Project.

We carried out pre-processing using Fieldtrip^32^ or custom written MATLAB (Version R2019a)^33^ scripts. Analysis was only undertaken on the following 19 EEG channels: Fp1, Fp2, Fz, F3, F7, F4, F8, T3, C3, Cz, C4, T4, T5, P3, Pz, P4, T6, O1, O2 (channels labelled T7, T8, P7, P8 were used for Danish EEGs as are in the same location as T3, T4, T5, T6 in the traditional 10-20 system). Any additional channels were removed. Data were re-referenced to average montage and resampled to the minimum sampling rate of all data (256Hz). EEGs were then segmented at the pre-defined epochs (10-20s eyes-closed, resting-state sections). Epochs across all three subgroups ((i) BIOJUME (JME); (ii) LEAP controls and (iii) Chowdhury controls) were independently reviewed and confirmed to ensure consistency in epoch choice between subgroups.

### EEG features

Twelve measures (six frequency-based and six network-based) were extracted from JME and control EEGs. Full methodology for the calculation of EEG features is presented in Supplementary Methods but outlined briefly below.

#### Frequency-based

Independent component analysis (ICA) was used to remove any remaining artifacts existing in the EEG between 2-20Hz and re-referenced to average montage. We calculated the mean relative power spectral density (PSD) in (i) delta (2-5Hz), (ii) low-alpha (6-9Hz), (iii) high-alpha (10-11Hz) and (iv) beta (12-19Hz) frequency bands; (v) the peak alpha frequency (PAF) and (vi) the shift in alpha PSD from high to low-alpha (alpha shift).

#### Network-based

Functional networks were inferred using phase-locking value (PLV) on EEG epochs Butterworth bandpass filtered between 6-9Hz. Five measures were used to characterise functional network topology: (i) mean strength, (ii) mean strength variance, (iii) average clustering coefficient, (iv) average characteristic path length and (v) small-world index. Additionally, the propensity of networks to generate seizure-like dynamics was modelled using the framework of Brain Network Ictogenicity (BNI).^24, 25^ Larger BNI values indicate that the brain network has a higher tendency to transmit from normal to seizure-like dynamics.

### Statistical Analysis Procedure

Statistical analysis was carried out in SPSS^34^ software and graphics produced in GraphPad Prism. Prior to statistical testing, violations of test assumptions were checked, and statistical tests chosen accordingly. Transformations and removal of outliers were done where required. Hedge’s g was used to calculate effect sizes.

#### Demographics, clinical characteristics and EEG details

Differences in age, epoch length and EEG time between cohorts were testing using two-tailed Mann-Whitney tests. Differences in sex distribution were tested using two-tailed chi-squared tests.

#### Influence of confounding factors on EEG features

Analysis of associations between potential confounders and EEG features were tested using Spearman’s rank tests for continuous variables (age, EEG time, epoch length) and two-tailed Mann-Whitney tests (stratified by JME and control groups) for sex. For analysis of EEG time, 11 JME EEGs with recording times between 23.35 and 00.21 were excluded, leaving only EEGs with recording times between 8:00-20:00. Differences in EEG features between JME individuals not on ASM therapy (untreated) and those on ASM therapy (treated) at the time of the EEG were tested using multiple linear regression models of each EEG feature including age as a covariate. The same method was also used to test for differences in EEG features between BIOJUME recruitment sites.

#### EEG features in JME compared to controls

Disparities in EEG features in JME and controls were first tested univariately using Mann-Whitney tests. P-values were corrected for multiple comparisons for the four frequency bands in PSD analysis and for the five measures of network topology using Bonferroni-holm. Kruskal Wallis tests were then used to test for differences in EEG features between the two control groups (LEAP & Chowdhury controls) and JME, again corrected for multiple comparisons for the four frequency bands and for the five measures of network topology using Bonferroni-holm. If these tests were significant (corrected p-value <0.05), Bonferroni-holm corrected Mann-Whitney tests were carried out between each of the three groups.

Using the results of the above analysis, multiple linear regression analysis was performed for each EEG feature to investigate differences between JME and control groups whilst accounting for the appropriate covariates (sex, age, epoch length, EEG time, ASM treatment, control subgroup). For EEG features where there was an opposing direction of effect between the two control groups and JME, LEAP controls and Chowdhury controls were added separately into the regression models as dummy variables (JME=0/LEAP=1; JME=0/Chowdhury=1). This enabled differences between each control subgroup to be detected rather than combining the two control groups. Similarly, for EEG measures which differed between untreated and treated JME EEGs, these were also added separately to the model as dummy variables (control=0/untreated JME=1; control=0/treated JME=1). Estimated marginal means of EEG measures for subgroups were obtained from these multivariable regression models.

#### ROC analysis

For those EEG features which were significantly different in univariate JME vs control analysis, receiver operating characteristic (ROC) curves were calculated to quantify their ability to discriminate JME and control EEGs. An optimal cut-off point was obtained for the EEG feature with the highest ROC area under the curve (AUC), using the value with the highest sensitivity and specificity values. Positive and negative predictive values were then calculated using standard methods.^35^

#### Heterogeneity of EEG features within the JME cohort

Multiple linear regression analysis was performed to test for associations of EEG features with clinical variables (seizure type, seizure control, PPR) within the JME cohort. Estimated marginal means of EEG measures for JME subgroups were obtained from these multivariable regression models.

#### Test re-test reliability of EEG measures

Two methods were used to investigate the test-retest reliability of each EEG measure: (i) Spearman’s rank correlation coefficient and (ii) intra-class correlation (ICC). The test-retest reliability of EEG features were tested first between epochs within the same EEG recording session (*between-epoch*) and second, between EEGs taken at different recording sessions but in the same ASM treatment state (both EEG taken whilst untreated or both while treated) (*between-EEG*). For the between-epoch analysis a two-way mixed model ICC with absolute agreement was used, using single measures. For the between-EEG analyses a two-way mixed model ICC with absolute agreement, using average measures was used, as here, the reliability of average measures taken at the two EEG recordings was being tested. To control for the effect of epoch length, we ran further analysis comparing EEG measures between epochs of the same/ similar (±5s) length.

### Ethical approval

BIOJUME is funded by the Canadian Institutes of Health Research (MOP-142405) and received ethical approval from the National Health Service (NHS) Health Research Authority (South Central - Oxford C Research Ethics Committee, reference 16/SC/0266) and the Research Ethics Board of the Hospital for Sick Children, Toronto (REB#1000033784). Local ethical approvals were also held for all international sites. All procedures complied with appropriate regulatory requirements and ethical principles in line with the Declaration of Helsinki. Informed consent was obtained and documented for all participants. Assent was obtained from minors (under 16), and informed consent was obtained on their behalf by a parent or legally appropriate guardian. All data from participants were de-identified before entry onto the central database.

### Data availability

The data supporting the findings of this study are available from the corresponding author upon reasonable request.

## Results

### Demographics, clinical characteristics and EEG details

194 EEGs from 147 individuals with JME, and 95 EEGs from 95 control participants were included in this study. Demographics from both groups and EEG details are presented in Table 2, with the JME group further stratified by ASM treatment status and the control group broken down by data origin. There were significant age (U=5156, p=0.003) and sex (χ^2^=7.3, p=0.007) differences between JME and control cohorts. Further, EEG time was significantly different between JME and control EEGs, with JME EEGs occurring earlier than control EEGs (U=661, p=2.6×10^-13^). Whilst all EEG epochs were between 10-20 seconds long, control epochs were on average significantly longer than JME epochs (U=4353, p=1.7×10^-8^). The clinical characteristics of the JME cohort are presented in Table 3.

**Table 2.**
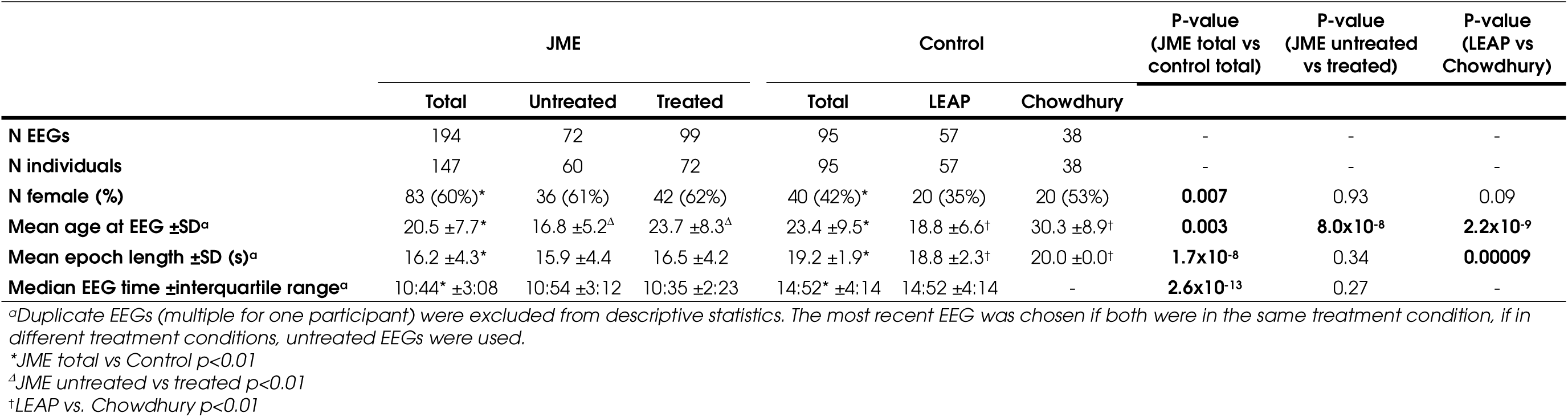
Demographics of JME and control participants included in the EEG study. P-values for continuous dependent variables are from Mann-Whitney tests and for categorical variables from Chi-squared tests. JME = Juvenile Myoclonic Epilepsy.

**Table 3.**
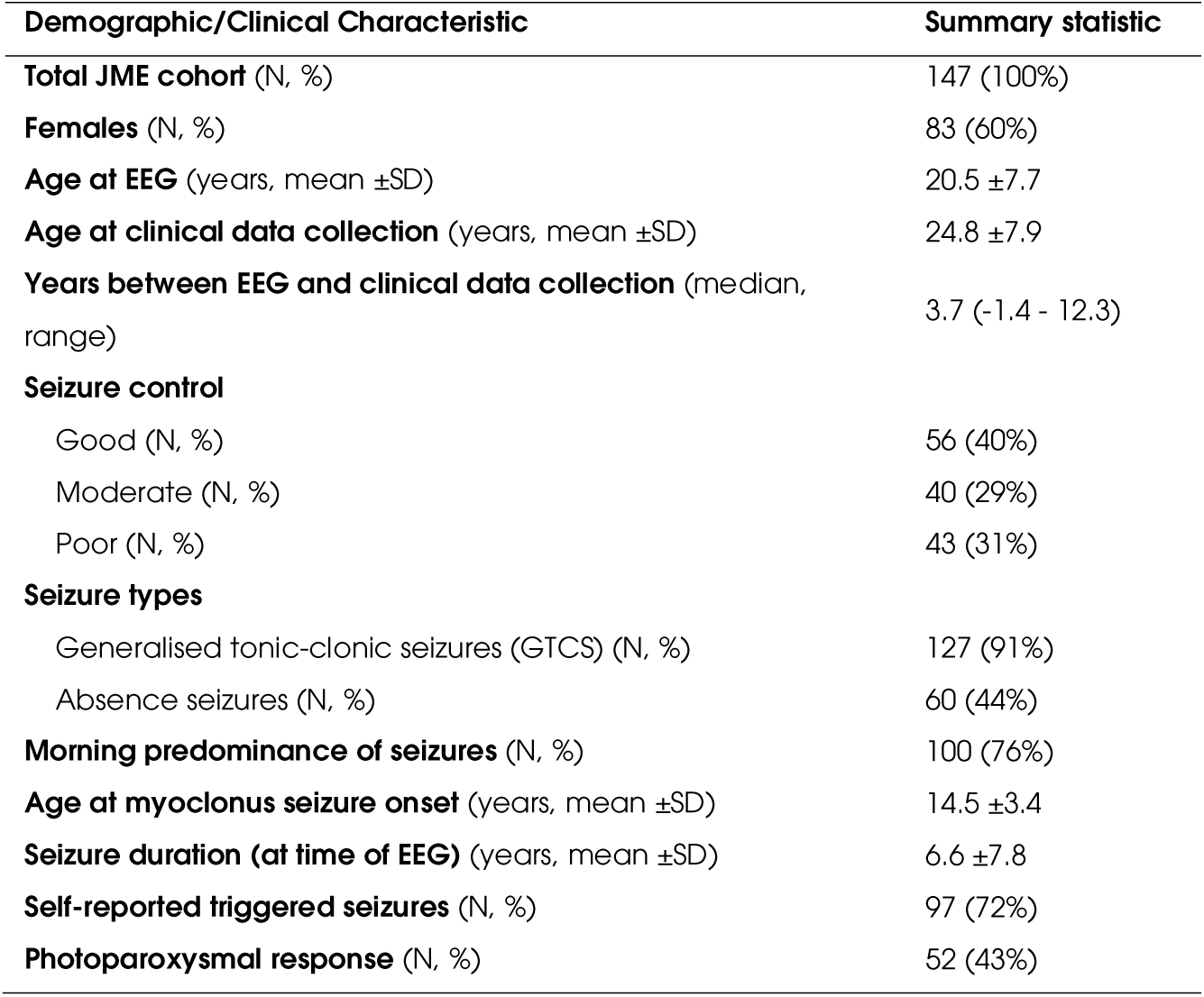
Demographics and clinical characteristics of participants included in this study. Percentage denominators are adjusted for missing data. JME = Juvenile Myoclonic Epilepsy.

### Influence of confounding factors on EEG features

#### Sex

There was a weak association of higher PAF in females with JME (U=1841, p=0.07), otherwise there were no sex differences in any EEG feature in JME or control groups.

#### Age, epoch length and EEG time

Figure 2A shows the results of Spearman’s rank correlation between each EEG feature and potentially confounding continuous variables (age, EEG time and epoch length) in controls and JME. Note that information on EEG time was only available for LEAP controls and JME, not Chowdhury controls. Age and epoch length were associated with a variety of frequency-based and network-based EEG features, whilst EEG time was associated only with frequency-based features. Interestingly, the relationship between PAF and EEG time was opposing in controls (r_s_=-0.38, p=0.015), and JME (r_s_=0.18, p=0.038).

**Figure 2.**
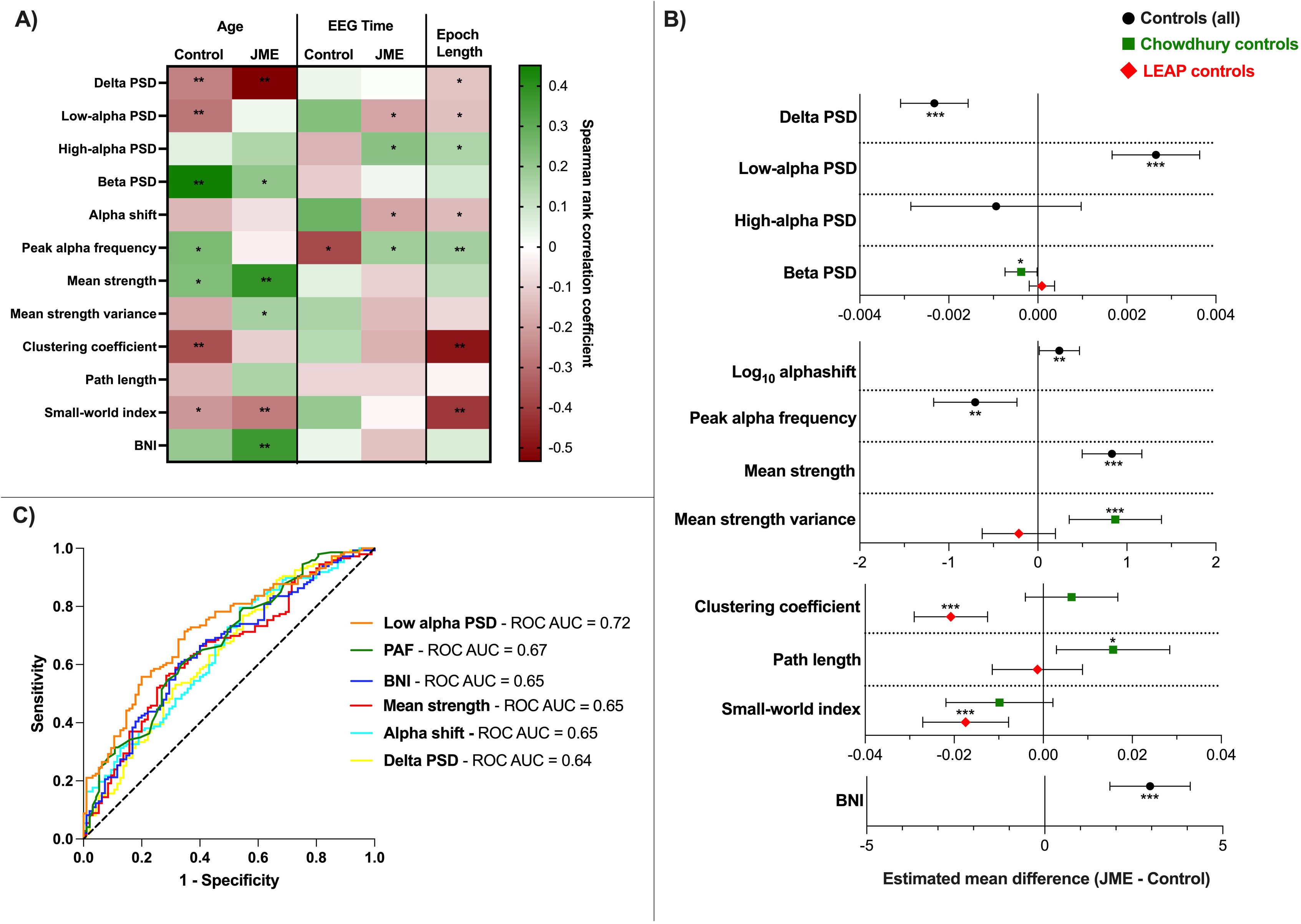
**A)** A heatmap representing the influence of continuous covariates on each EEG feature. Colour represents the Spearman’s rank correlation coefficient between the continuous covariates and EEG features (green = positive correlation, red = negative correlation). Age and EEG time results are stratified by JME/Controls. *p<0.05, **p<0.01. **B)** A summary of the estimated marginal mean difference of EEG features in JME compared to controls, from multiple linear regression models controlling for epoch length, age (for all measures) and EEG time (for log_10_ alpha shift and PAF only). The central marker shows estimated marginal mean difference and error bars are 95% confidence intervals. *p<0.05, **p<0.01 and ***p<0.001 in multiple linear regression model. **C)** ROC curves for EEG features in JME and controls, with area under the ROC AUC values in the legend. BNI = Brain Network Ictogenicity; JME = Juvenile Myoclonic Epilepsy; LEAP = Longitudinal European Autism Project; PAF = Peak alpha frequency; PSD = Power spectral density; ROC AUC = Receiver operating curve, area under the curve.

#### ASM treatment

Low-alpha PSD was significantly higher in untreated JME EEGs compared to treated JME EEGs (b=-0.002, p=0.03), whilst controlling for age. P-values for ASM treatment status were >=0.1 for all other EEG features.

#### EEG site

The site at which BIOUME EEGs were recorded had no significant association with any frequency-based (p>0.28) EEG features. Clustering coefficient was the only feature to show any differences between sites (p=0.053, Figure S 3). No other network-based measure significantly differed between sites (p>0.20).

### EEG measures in JME compared to controls

Table 4 shows the results of univariate tests of EEG features between controls and JME. Average relative PSD plots for both JME and controls are displayed in Figure S 3. Relative delta PSD and PAF are significantly lower in JME than controls whilst relative low-alpha PSD, alpha shift, mean strength and BNI are significantly higher in JME in univariate analysis. Relative low-alpha PSD has the greatest effect size of g=0.72. Analysis with stratified control subgroups (Table 4) shows an opposite direction of effect in LEAP and Chowdhury control subgroups compared to JME for relative beta PSD, mean strength variance, average clustering coefficient, average characteristic path length and small-world index. To account for different subgroups and the effect of confounders discussed above, multiple linear regression models were performed for each EEG measure, containing the appropriate covariates (age, sex, epoch length, EEG time). A summary of the differences in EEG features between JME and Controls are presented in Figure 2B and all regression models presented in Supplementary Material Tables S2-6. Relative low-alpha PSD (untreated JME b=0.0033, p=2.9×10^-7^; treated JME b=0.0022, p=9.1×10^-5^) and BNI (b=3.0, p=5.1×10^-7^) were the features with the greatest difference between JME and controls whilst accounting for covariates (Figure 2B).

**Table 4.** Summary of p-values from univariate statistical tests on EEG features between groups. BNI = Brain Network Ictogenicity; JME = Juvenile Myoclonic Epilepsy; LEAP = Longitudinal European Autism Project; PSD = Power spectral density; ROC AUC = Receiver operating curve, area under the curve.

| EEG feature | Control vs. JME |  |  |  | Stratified control groups |  |  |  |
| --- | --- | --- | --- | --- | --- | --- | --- | --- |
|  | Uncorrected p-values | Bonferroni-holm corrected p-values | Hedge's g | ROC AUC | Comparison | Mean diff | Uncorrected p-values | Bonferroni-holm corrected p-values |
| Relative Delta PSD | **0.00024** | **0.00072** | 0.55 | 0.64 | Difference between groups (Kruskal-Wallis) | - | **0.0011** | **0.0023** |
|  |  |  |  |  | LEAP vs. JME (Mann-Whitney) | 0.0014 | **0.0024** | **0.0072** |
|  |  |  |  |  | Chowdhury vs. JME (Mann-Whitney) | 0.0020 | **0.0064** | **0.013** |
|  |  |  |  |  | LEAP vs. Chowdhury (Mann-Whitney) | -0.00063 | 0.64 | 0.64 |
| Relative Low-alpha PSD | **7.2x10<sup>-9</sup>** | **2.9x10<sup>-8</sup>** | 0.80 | 0.72 | Difference between groups (Kruskal-Wallis) | - | **6.7x10<sup>-9</sup>** | **2.7x10<sup>-8</sup>** |
|  |  |  |  |  | LEAP vs. JME (Mann-Whitney) | -0.0023 | **0.00008** | **0.00016** |
|  |  |  |  |  | Chowdhury vs. JME (Mann-Whitney) | -0.0035 | **7.2x10<sup>-8</sup>** | **2.2x10<sup>-7</sup>** |
|  |  |  |  |  | LEAP vs. Chowdhury (Mann-Whitney) | 0.0012 | **0.013** | **0.013** |
| Relative High-alpha PSD | 0.10 | 0.20 | 0.29 | - | Difference between groups (Kruskal-Wallis) | - | 0.13 | 0.13 |
|  |  |  |  |  | LEAP vs. JME (Mann-Whitney) | 0.0024 | **0.042** | 0.13 |
|  |  |  |  |  | Chowdhury vs. JME (Mann-Whitney) | 0.0010 | 0.73 | 0.73 |
|  |  |  |  |  | LEAP vs. Chowdhury (Mann-Whitney) | 0.0014 | 0.26 | 0.52 |
| Relative Beta PSD | 0.38 | 0.38 | 0.22 | - | Difference between groups (Kruskal-Wallis) | - | **2.5x10<sup>-5</sup>** | **7.5x10<sup>-5</sup>** |
|  |  |  |  |  | LEAP vs. JME (Mann-Whitney) | -0.00014 | 0.083 | 0.083 |
|  |  |  |  |  | Chowdhury vs. JME (Mann-Whitney) | 0.00060 | **0.00013** | **0.0003** |
|  |  |  |  |  | LEAP vs. Chowdhury (Mann-Whitney) | -0.00073 | **1.3x10<sup>-5</sup>** | **3.9x10<sup>-5</sup>** |
| Peak alpha frequency | **1.3x10<sup>-5</sup>** | - | 0.66 | 0.67 | Difference between groups (Kruskal-Wallis) | - | **1.4x10<sup>-5</sup>** | - |
|  |  |  |  |  | LEAP vs. JME (Mann-Whitney) | 0.45 | **0.0044** | **0.0088** |
|  |  |  |  |  | Chowdhury vs. JME (Mann-Whitney) | 0.84 | **2.1x10<sup>-5</sup>** | **6.3x10<sup>-5</sup>** |
|  |  |  |  |  | LEAP vs. Chowdhury (Mann-Whitney) | -0.39 | **0.039** | **0.039** |
| Log<sub>10</sub> alpha shift | **0.00012** | - | 0.54 | 0.65 | Difference between groups (Kruskal-Wallis) | - | **0.00059** | - |
|  |  |  |  |  | LEAP vs. JME (Mann-Whitney) | -0.21 | **0.0016** | **0.0048** |
|  |  |  |  |  | Chowdhury vs. JME (Mann-Whitney) | -0.24 | **0.0037** | **0.0074** |
|  |  |  |  |  | LEAP vs. Chowdhury (Mann-Whitney) | 0.030 | 0.77 | 0.77 |
| Mean strength | **0.00012** | **0.0006** | 0.48 | 0.65 | Difference between groups (Kruskal-Wallis) | - | **0.00022** | **0.00066** |
|  |  |  |  |  | LEAP vs. JME (Mann-Whitney) | -0.73 | **0.00013** | **0.00039** |
|  |  |  |  |  | Chowdhury vs. JME (Mann-Whitney) | -0.38 | **0.039** | 0.078 |
|  |  |  |  |  | LEAP vs. Chowdhury (Mann-Whitney) | -0.35 | 0.067 | 0.078 |
| Mean strength variance | 0.15 | 0.52 | 0.22 | - | Difference between groups (Kruskal-Wallis) | - | **9.0x10<sup>-5</sup>** | **0.00037** |
|  |  |  |  |  | LEAP vs. JME (Mann-Whitney) | 0.063 | 0.32 | 0.32 |
|  |  |  |  |  | Chowdhury vs. JME (Mann-Whitney) | -0.80 | **0.00012** | **0.00024** |
|  |  |  |  |  | LEAP vs. Chowdhury (Mann-Whitney) | 0.86 | **5.6x10<sup>-5</sup>** | **0.00017** |
| Clustering coefficient | 0.33 | 0.66 | 0.10 | - | Difference between groups (Kruskal-Wallis) | - | **2.8x10<sup>-7</sup>** | **1.4x10<sup>-6</sup>** |
|  |  |  |  |  | LEAP vs. JME (Mann-Whitney) | 0.011 | **0.033** | **0.033** |
|  |  |  |  |  | Chowdhury vs. JME (Mann-Whitney) | -0.023 | **9.0x10<sup>-6</sup>** | **1.8x10<sup>-5</sup>** |
|  |  |  |  |  | LEAP vs. Chowdhury (Mann-Whitney) | 0.034 | **1.0x10<sup>-7</sup>** | **3.0x10<sup>-7</sup>** |
| Path length | 0.13 | 0.52 | 0.17 | - | Difference between groups (Kruskal-Wallis) | - | **0.022** | **0.044** |
|  |  |  |  |  | LEAP vs. JME (Mann-Whitney) | 0.00059 | 1.0 | 1.0 |
|  |  |  |  |  | Chowdhury vs. JME (Mann-Whitney) | -0.014 | **0.0071** | **0.021** |
|  |  |  |  |  | LEAP vs. Chowdhury (Mann-Whitney) | 0.014 | **0.022** | **0.044** |
| Small-world index | 0.61 | 0.66 | 0.02 | - | Difference between groups (Kruskal-Wallis) | - | **0.040** | **0.044** |
|  |  |  |  |  | LEAP vs. JME (Mann-Whitney) | 0.0080 | 0.074 | 0.15 |
|  |  |  |  |  | Chowdhury vs. JME (Mann-Whitney) | -0.0093 | 0.19 | 0.19 |
|  |  |  |  |  | LEAP vs. Chowdhury (Mann-Whitney) | 0.017 | **0.013** | **0.039** |
| BNI | **5.6x10<sup>-5</sup>** | - | 0.56 | 0.65 | Difference between groups (Kruskal-Wallis) | - | **0.00021** | - |
|  |  |  |  |  | LEAP vs. JME (Mann-Whitney) | -2.87 | **0.00021** | **0.00063** |
|  |  |  |  |  | Chowdhury vs. JME (Mann-Whitney) | -1.47 | **0.012** | **0.024** |
|  |  |  |  |  | LEAP vs. Chowdhury (Mann-Whitney) | -1.40 | 0.19 | 0.19 |

### ROC analysis

To test which EEG feature best served to discriminate JME and control EEGs, ROC curve analysis was performed for delta PSD, low-alpha PSD, PAF, alpha shift, mean strength and BNI, based on the results of the analysis above. Low-alpha PSD had the greatest ROC AUC of 0.72 (95% CI=0.66-0.79, p=7.2×10^-9^), followed by PAF (AUC=0.67, 95% CI=0.60-0.74, p=7.4×10^-6^) and BNI (AUC=0.65, 95% CI=0.58-0.72, p=0.000056) (Figure 2C).

A proposed cut-off value for low-alpha PSD is >0.0066, which has a sensitivity of 0.69 and a specificity of 0.67 (Figure S 2). Using this threshold of relative low-alpha PSD gives a positive predictive value (PPV) of 77% and a negative predictive value (NPV) of 58%.

Using only untreated JME EEGs compared to controls increases the ROC AUC for low-alpha PSD to 0.78 (95% CI=0.70-0.85, p=6.2×10^-9^). The same thresholding method gave the same cut-off value of >0.0066, as above (sensitivity=75%, specificity=67%), giving a PPV of 59% and a NPV of 81%.

### Heterogeneity of EEG features within JME cohort

Figure 3 shows a summary of associations of each EEG feature with a variety of clinical variables in the JME cohort, showing standardized beta coefficients from multiple linear regression models controlling for age and epoch length. The greatest standardized beta coefficient changes from 0 exist for functional network topology measures, particularly average clustering coefficient, average characteristic path length and small-world index. All clinical outcomes in Figure 3 (absence seizures, lack of PPR, no morning predominance of seizures and triggered seizures) have been associated with worse seizure control in this dataset^36^.

**Figure 3.**
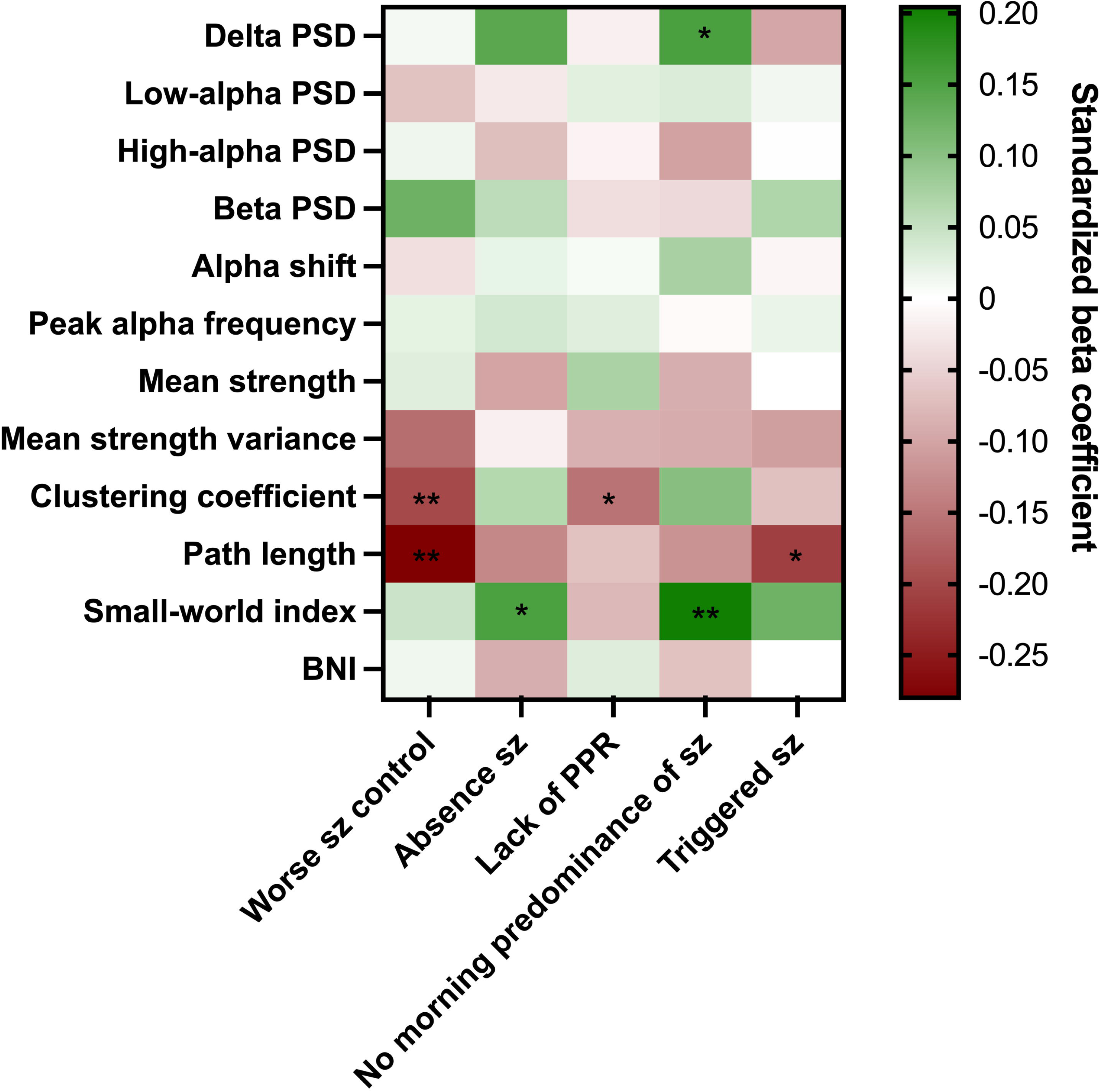
Heatmap showing the standardized beta coefficients of clinical variables in multiple linear regression models of EEG features, controlling for age and epoch length. *p<0.05, *p<0.01. BNI = Brain Network Ictogenicity; PPR = photoparoxysmal response; PSD = Power spectral density; Sz = seizures.

### Test-retest reliability

Table 5A shows the test-retest reliability between epochs in the same EEG recording and Table 5B between multiple EEG recordings in the same ASM treatment state in JME participants. Between epochs, there is excellent reliability (r_s_>0.87, ICC>0.88) for all frequency-based measures, with differences in epoch length having a minimal effect on reliability. However, for network topology measures there is lower test-retest reliability but improving when epoch length is consistent. BNI has a good reliability (r_s_ >0.73, ICC>0.77), with epoch length having little effect.

**Table 5.** A) The test-retest reliability of EEG features in different epochs from the same EEG. ICC (intra-class correlation). B) The test-retest reliability of EEG features in multiple EEG recordings taken when the participant was in the same treatment state (either both in the untreated state or both in the treated state). BNI = Brain Network Ictogenicity; JME = Juvenile Myoclonic Epilepsy; PSD = Power spectral density.

| A) Test-retest reliability (between epochs in same EEG) |  |  |  |  |
| --- | --- | --- | --- | --- |
| EEG feature | Spearman's r |  | ICC |  |
|  | All epochs (n=178) | Equal length epochs (n=116) | All epochs (95% CI) (n=178) | Equal length epochs (n=116) |
| Delta PSD | 0.90 | 0.90 | 0.91 (0.88-0.93) | 0.91 (0.88-0.94) |
| Low-alpha PSD | 0.90 | 0.92 | 0.91 (0.88-0.93) | 0.93 (0.91-0.95) |
| High-alpha PSD | 0.90 | 0.91 | 0.88 (0.84-0.91) | 0.91 (0.87-0.94) |
| Beta PSD | 0.91 | 0.95 | 0.90 (0.87-0.92) | 0.94 (0.92-0.96) |
| Peak alpha frequency | 0.88 | 0.86 | 0.87 (0.84-0.91) | 0.89 (0.85-0.92) |
| Log<sub>10</sub> alpha shift | 0.91 | 0.91 | 0.91 (0.88-0.93) | 0.93 (0.90-0.95) |
| Mean strength | 0.79 | 0.83 | 0.82 (0.76-0.86) | 0.85 (0.79-0.89) |
| Mean strength variance | 0.69 | 0.74 | 0.73 (0.66-0.80) | 0.78 (0.70-0.84) |
| Clustering coefficient | 0.41 | 0.36 | 0.59 (0.48-0.68) | 0.64 (0.52-0.74) |
| Path length | 0.45 | 0.54 | 0.49 (0.38-0.60) | 0.55 (0.41-0.67) |
| Small-world index | 0.44 | 0.44 | 0.55 (0.44-0.64) | 0.58 (0.45-0.69) |
| BNI | 0.76 | 0.81 | 0.78 (0.72-0.83) | 0.81 (0.73-0.86) |

**B) Test-retest reliability (between multiple EEGs in same treatment state)**
| EEG feature | Spearman's r |  | ICC |  |
| --- | --- | --- | --- | --- |
| | All epochs<br>(n=21) | Equal length epochs $\pm 5s$<br>(n=13) | All epochs (95% CI)<br>(n=21) | Equal length epochs $\pm 5s$<br>(n=13) |
| Delta PSD | 0.70 | 0.80 | 0.83 (0.57-0.93) | 0.86 (0.54-0.96) |
| Low-alpha PSD | 0.72 | 0.59 | 0.78 (0.48-0.91) | 0.73 (0.09-0.92) |
| High-alpha PSD | 0.34 | 0.45 | 0.51 (-0.24-0.81) | 0.55 (-0.58-0.86) |
| Beta PSD | 0.72 | 0.70 | 0.87 (0.68-0.95) | 0.89 (0.62-0.97) |
| Peak alpha frequency | 0.34 | 0.18 | 0.48 (-0.27-0.79) | 0.36 (-1.34-0.81) |
| Log<sub>10</sub> alpha shift | 0.54 | 0.43 | 0.67 (0.22-0.87) | 0.62 (-0.34-0.89) |
| Mean strength | 0.74 | 0.67 | 0.85 (0.65-0.94) | 0.84 (0.51-0.95) |
| Mean strength variance | 0.38 | 0.03 | 0.67 (0.18-0.87) | 0.55 (-0.59-0.86) |
| Clustering coefficient | 0.18 | 0.41 | 0.54 (-0.05-0.81) | 0.86 (0.51-0.96) |
| Path length | 0.42 | 0.42 | 0.78 (0.45-0.91) | 0.77 (0.25-0.93) |
| Small-world index | 0.46 | 0.63 | 0.72 (0.29-0.89) | 0.86 (0.53-0.96) |
| BNI | 0.64 | 0.55 | 0.80 (0.50-0.92) | 0.75 (0.24-0.92) |

The results presented in Table 5B should be interpreted with more caution due to the smaller number of EEGs available for this analysis (total n=21) and the potential effect of age on these measures (13 EEGs were performed <=1 year apart, 8 EEGs between 2-10 years apart). To account for epoch length, we further ran the analysis including only epochs ±5s difference in length. The reliability is much more variable across the frequency and network topology measures, however, remains good (ICC>0.75) for delta, low-alpha and beta PSD, mean strength, path length and BNI.

## Discussion

This study uses a range of EEG features to characterise differences in the interictal EEG of individuals with JME compared to controls and assess their heterogeneity within these cohorts. Our results support findings from previous studies in IGE,^5, 7, 17^ showing that the low-alpha band is the frequency range which is the most abnormal, with significantly elevated low-alpha PSD and lower peak alpha frequency in JME compared to controls. Further, we show that these findings are reproducible when using EEGs from multiple sites and multiple cohorts of patients, giving this finding considerable external validity and potential for application in settings outside of research. A novel finding was increased BNI in functional networks derived from JME EEGs, which corroborates the same findings in other epilepsies^18, 24, 25^ and using other modalities, such as MEG.^27^

Functional network topology measures, such as average clustering coefficient, average characteristic path length and small-word index (which is derived from the prior two measures), were the only features which consistently differ according to JME phenotype. JME patients with poor seizure control (or risk factors for it) have significantly lower normalized path length and normalized clustering coefficient than those with good seizure control.

In addition, our results highlight that confounding factors such as age and epoch length have a considerable effect on many EEG features and so should be carefully controlled for. These confounding factors may explain the variability of findings from previous studies investigating network topology measures in epilepsy.

### Low-alpha oscillatory activity as a potential biomarker of JME

ROC analysis showed that low-alpha PSD has ‘acceptable’ clinical discriminatory ability (AUC between 0.7-0.8),^37^ with PAF and BNI falling just below this threshold at 0.67 and 0.65 respectively. However, p-values associated with these tests show these measures as highly significant in their ability to differentiate above chance. Further investigation of relative low-alpha PSD as a biomarker showed satisfactory sensitivity (69%), specificity (67%), PPV (77%) and NPV (58%) in analysis including all JME individuals, with biomarker quality improving when comparing controls to only untreated JME individuals (AUC improvement from 0.72 to 0.78). This suggests low-alpha PSD may have promise as a biomarker of JME, however, this requires replication in independent cohorts. A previous study by Schmidt et al. (2016)^19^ investigated EEG biomarkers of IGE and found that a local coupling biomarker best classified IGEs from controls, compared to other EEG parameters such as average power across the whole EEG power spectrum, peak alpha frequency and mean degree of functional networks. However, this study did not investigate the low-alpha PSD specifically and investigated all IGE syndromes, not just JME.

### Significance of the alpha frequency in epilepsy

Neural oscillations in the alpha frequency band are thought to arise from cortico-thalamic interactions and govern top-down control of cognitive processes and feedback processing from higher-order cortical areas to lower-order visual areas.^10^ However, the exact mechanisms of alpha oscillations are complex and still poorly understood. Despite the mechanistic complexity, it is clear that alterations in alpha oscillations are seen in a number of neurological and psychiatric disorders, such as dementia, schizophrenia, stroke^38^ and epilepsy. The change in alpha oscillatory activity from a higher frequency in healthy controls to a lower frequency in epilepsy patients, is of particular relevance and interest to IGE syndromes, since dysfunction in thalamocortical connections are thought to drive the generation of spike-wave discharges and generalised seizures.^11, 12^

Unlike previous work by Abela et al. (2019),^5^ we observed no significant difference in EEG features relating to alpha oscillatory activity (low-alpha PSD, peak alpha frequency and alpha shift) between any phenotypic group in this cohort. This agrees with recent evidence presented by Pegg et al. (2020)^7^ which showed no difference in PSD between well-controlled and drug-resistant patients with IGE. In addition, and in agreement with Pegg et al. (2020),^7^ no other PSD measures in other frequency bands showed associations with any clinical features or prognostic outcomes in JME.

In the present study, EEGs were obtained from patients at any point during their JME clinical care, not necessarily at the time of recruitment, with a median of 4 years difference between EEG acquisition and recruitment. Hence, their reported seizure control at the time of recruitment may not accurately reflect their seizure control at the time the EEG was taken. Because of this, the present study aimed to identify differences in resting-state EEG activity that are not dependent on current seizure control but rather state-independent, such as a susceptibility drug resistance, or a history of specific seizure types. Other studies of this kind, such as Abela et al. (2019)^5^ and Pegg et al. (2020)^7^ recorded EEGs at the time of recruitment, and therefore were investigating differences in EEG activity based on current seizure status. Definitions of seizure control also differ between studies, with the present study and Pegg et al. (2020)^7^ using the commonly used definition of drug resistance in epilepsy^39^ (continuation of seizures despite at least 2 ASM trials), whereas Abela et al. (2019)^5^ defined those with poor seizure control as four or more seizures of any type during the 12 months prior to the study inclusion.

### Network topology measures

Our results indicate that functional network topology, as derived from activity in the 6-9Hz frequency range, is inherently different in JME compared to healthy controls, supporting evidence from other studies in IGE.^17, 19–22, 40^ We can conclude that these differences are not due to ASM treatment as there were no differences in network topology measures between untreated and treated individuals with JME. The mean strength of JME functional networks was significantly higher than controls in both univariate and multivariable analysis, and the variance of mean strength was also significantly higher in JME than Chowdhury controls, replicating results from the original study.^17^ An increase in the mean strength of networks in JME indicates that the EEG signal, and therefore brain oscillatory activity, at each node is synchronised to the activity at other nodes to a higher extent than in controls. However, the increase in mean strength variability also indicates that there may be both abnormally under-connected brain regions as well as over-connected regions. The opposing direction of effect between clustering coefficient and path length in Chowdhury/LEAP control networks compared to JME is unexpected and puzzling, and potential factors influencing this, including differences in age and epoch length, are discussed below.

Alterations in functional network topology also showed associations with clinical features and outcomes in JME, particularly clustering coefficient, which reflects a network’s functional segregation, and path length, which reflects a network’s functional integration. Shorter average path length and decreased clustering coefficient were associated with poorer seizure control in this JME cohort. Shorter path length was also associated with experiencing triggered seizures, and decreased clustering coefficient was associated with *not* experiencing PPR, both phenotypes associated with having a worse seizure outcome in this cohort.^36^ A short average path length and low clustering is representative of more random networks, whereby information can pass easily through the network from one node to functionally distinct nodes due to longer range functional connections,^41^ and therefore, speculatively, may have an increased likelihood to synchronize more easily, implying an increased vulnerability to seizures. Indeed, JME networks have been shown to transition to more random network topology during spike-wave discharges, with decreased clustering in theta and beta frequency bands.^42^

Conversely, higher small-world index, a measure reflecting the small-worldness of the networks derived from CC and PL, was significantly associated with experiencing absence seizures. A higher small-world index indicates networks have more regular, lattice-like structures, with high clustering, but also with long range connections from functionally distinct regions, keeping the average path length short. Given that experiencing absence seizures is strongly associated with poorer seizure outcome,^43^ it is somewhat surprising that differing network types (random vs. ordered) are associated with these two phenotypes. However several studies of network topology in patients with absence seizures have also shown increased clustering, both in the ictal^44^ and interictal state,^40^ indicating networks with a more ordered, lattice-like topology may be more vulnerable to absence seizures. In addition, a study by Lee et al. (2020)^22^ showed that individuals with absence epilepsy had the highest small-world index compared to other IGE syndromes, including JME.

Previous literature on functional connectivity in epilepsy is variable, since results can differ depending on modality (EEG, MEG, fMRI), functional connectivity measure (phase-locking value, coherence, correlation etc.) and the frequency band (low-alpha (6-9Hz), high-alpha (10-11Hz) or wider bands (2-20Hz)), therefore making comparisons between the present results and previous evidence challenging. A study from Pegg et al. (2021),^21^ using EEG derived functional networks from patients with IGE, showed no differences in network topology between well-controlled (n=19) and drug-resistant patients (n=18) at 6-9Hz nor 10-11Hz, using phase-locking value. Potential reasons for this difference in results include sampling differences (IGE vs. JME only) and a small sample size in the prior study,^21^ perhaps leading to a lack of statistical power to detect the differences apparent in this larger cohort.

### Dynamic Network Models

As well as differences in static network topology, this study also investigated differences in JME functional networks using Brain Network Ictogenicity, a computational framework that uses dynamic model to characterise the likelihood of brain networks to enter a seizure-like state. The use of dynamic models in epilepsy research has grown in popularity and BNI has shown promise both as a diagnostic and prognostic biomarker,^18, 19, 25, 27^ but also in understanding and modelling the transition of the brain into a seizure state. As hypothesised, this study revealed that individuals with JME have an increased likelihood of brain networks to transition to a seizure state (higher BNI) compared to controls.

Somewhat surprisingly, there was no association of any clinical features or outcomes with BNI. One may theorise that those with worse seizure control would have a higher BNI, as presumably these brain networks would be more susceptible to seizures, however, this was not the case. As mentioned previously, this may be because patients’ EEGs were not necessarily from the time of recruitment when they reported their current seizure/treatment status. However, the association of increased BNI with age in JME, but not controls, is of interest due to the collinearity of age with JME disease duration, as well as the probable follow-up-bias in the JME cohort, meaning that older individuals are more likely to have increased disease severity.

### Heterogeneity of EEG phenotypes

We observed that epoch length was highly influential on network topology measures, particularly clustering coefficient and small-world index, as well as lower frequency PSD measures, as has been shown in previous studies.^45^ Sex had no influence on any EEG feature in either JME or controls. Age was associated with multiple EEG features, as expected from knowledge of EEG and brain network changes through development. For example, delta PSD decreased with age in both JME and control cohorts supporting the understanding that delta oscillations are more prominent in younger children than adults.^46^ Additionally, beta power increased with age in both cohorts, reflecting the known association of beta power with maturation during adolescence.^47^

A further factor which we chose to investigate was the influence of time of day on EEG features. A morning predominance of seizures is a hallmark of JME and transcranial magnetic stimulation studies show that cortical excitability is highest in the morning in patients.^48, 49^ Hence, the fact that certain EEG parameters differ in JME throughout the day may reflect the differing seizure susceptibility threshold. This was apparent for low and high-alpha PSD, and therefore reflected in the alpha shift which decreased throughout the day in JME, but showed no relationship with EEG time in controls. There was also a significant increase in peak alpha frequency throughout the day in JME. Comparing the change in EEG features with time of day in JME and controls proved difficult as most JME EEGs took place in the morning, compared to most control EEGs which generally took place in the afternoon. Moreover, only LEAP controls had information on EEG time, further limiting the comparison. Matching future studies on EEG time would allow this diurnal change in hyperexcitability in JME and the effect on EEG features to be investigated further.

### Methodological considerations and limitations

In this study, functional networks were modelled in the 6-9Hz frequency range only. We chose this band based on the results of our frequency analysis showing that the low-alpha range showed the most significant changes in JME compared to controls, but also on previous evidence from frequency and network studies in IGE showing similar results.^5, 17, 19, 50^

All analysis in this study was carried out in EEG sensor space, whereby network nodes represent activity at EEG channels, rather than source space, where network nodes represent the brain regions determined to generate EEG activity. 19 channel clinical EEGs, such as those used in this study, limit the ability for EEG signals to undergo source reconstruction to determine the brain regions thought to generate the signal. Future studies using high density EEG electrode caps should assess whether there are benefits to using EEG source space vs sensor space, and may provide insights into brain regions influencing differences in functional network topology.

### Conclusion

Individuals with JME have increased power of neural oscillatory activity at low-alpha frequencies, along with increased BNI compared to controls, supporting evidence from studies in other epilepsies with considerable external validity. There is encouraging evidence for the use of low-alpha PSD as a biomarker of JME but requires replication in an independent dataset. Functional network topology measures are variable and prone to confounding but show significant associations with clinical features and outcomes in JME.

## Data Availability

All data produced in the present study are available upon reasonable request to the authors.

## Abbreviations

ASM: Antiseizure medication
BIOJUME: Biology of Juvenile Myoclonic Epilepsy
BNI: Brain Network Ictogenicity
GTCS: Generalised tonic-clonic seizure
IGE: Idiopathic Generalised Epilepsy
JME: Juvenile Myoclonic Epilepsy
PPR: Photoparoxysmal response

## Acknowledgements

The authors acknowledge all research staff at BIOJUME recruitment sites, in particular, those aiding with EEG collection (full list in Appendix); the BIOJUME phenotyping panel: Christoph P. Beier, Khalid Hamandi, Guido Rubboli, Marte Syvertsen and Rhys Thomas; members of the EU-AIMS Longitudinal European Autism Project (LEAP) and Fahmida Chowdhury for collecting control EEG data. Figure 1 created with BioRender.com.

