## Supplement for "Heterogeneity of resting-state EEG features in juvenile myoclonic epilepsy and controls"

### **Supplementary Material**

#### **Supplementary methods**

##### **Power spectral density (PSD) and frequency measures**

All PSD and frequency analysis was carried out using Fieldtrip or custom written MATLAB scripts. To remove any remaining artifacts in the data between 2-20Hz (e.g. eye or cardiac artifacts), independent component analysis (ICA) was carried out using Fieldtrip. Where ICA components or channels with high amounts of noise were removed, data were re-referenced to average.

The MATLAB p-welch function (using a 1-second Hanning window with 50% overlap) was used to perform the Fast Fourier Transform to provide the PSD from each epoch. The frequency range 2-20Hz was selected as it encapsulates frequency bands commonly used in the literature, while avoiding low‐frequency drifts (<2 Hz), as well as high frequency muscle artefacts (>20 Hz). The PSD between 2-20Hz, at 0.1Hz increments, was then normalised against the total power of each EEG channel to give the relative power. The mean relative power across all channels was then calculated for each EEG.

Data-driven frequency bands from Shackman et al. (2014)^1^ were used (delta: 2-5Hz, alpha-low: 6-9Hz, alpha-high: 10-11Hz, beta: 12-19Hz, and gamma: >21Hz). The lower limit of the delta frequency range was set to 2Hz and the gamma frequency range was excluded due to our previous selection of data in the 2-20Hz range only. The mean relative power in each frequency band was then calculated.

###### **Alpha Shift**

The shift in alpha power from high to low-alpha was calculated following methods in Abela et al. (2019).^2^ Relative power in the low-alpha frequency band (6-9Hz) was divided by relative power in the high-alpha frequency band (10-11Hz) to give the alpha shift. To normalise the distribution of this variable the log_10_ alpha shift was used for analysis.

###### **Peak alpha frequency**

To extract the peak alpha frequency from each EEG power spectrum, the 1/f background noise from the power spectrum was first removed to ensure that only oscillatory activity was being measured, based on methodology from Haller et al. (2018).^3^ This involved using the robust regression function in MATLAB to fit a line to the power spectrum in semi-log space, but to avoid the alpha region (6-13Hz) when fitting the line so that the alpha oscillatory signal did not bias the fit. The background noise underneath the line was then removed from the spectrum. From this, the MATLAB ‘findpeaks’ function was used to find the largest peak in the remaining oscillatory spectrum between 2 to 20Hz. The frequency at which this peak existed was then identified as the peak alpha frequency for each individual epoch.

##### **Inferring functional networks from EEG**

Inferring of functional networks uses methods described in Lopes et al. (2019).^4^ To summarise, functional networks were calculated using phase-locking value (PLV)^5-9^ on EEG epochs which had been Butterworth bandpass filtered between 6-9Hz. PLV was chosen due to its ability to differentiate IGE from controls.^8,9^ Network nodes were electrode locations and PLV values as network edges/connectivity weights. For each pair of nodes $i$ and $j$,

${PLV}_{ij}= \frac{1}{N_{s}}\left| \sum_{k=1}^{N_{s}} e^{i\Delta\phi_{ij}(t_{k})} \right|$,

where $N_{s}$ is the number of samples, and $\Delta\phi_{ij}(t_{k})$ is the instantaneous phase difference between the signals recorded from electrodes $i$ and $j$ at time $t_{k}$, computed using the Hilbert transform. We also found the average phase-lag $\tau_{ij}$between the two signals,

$\tau_{ij}=arg\left( \sum_{k=1}^{N_{s}} e^{i\Delta\phi_{ij}(t_{k})} \right)$.

Nodes $i$ and $j$ were considered connected if ${PLV}_{ij}$>0 and $\tau_{ij}$>0. Non-zero time-lag PLV was used to avoid false signal connections due to volume conduction.^10^ 99 surrogate networks were generated from the original EEG signals using the iterative amplitude-adjusted Fourier transform with 10 iterations.^11,12^ Connections were replaced with 0 if their ${PLV}_{ij}$ weights were below the 95% significance level compared to the same connection weights as computed from the surrogates. This method gave a directed, weighted functional network $a_{ij}$ from each EEG epoch.

**Network topology measures**

The methodology used to characterise the functional networks was adapted from Chowdhury et al. (2014)^13^ to account for our networks being weighted and directed. We used Brain Connectivity Toolbox (<http://www.brain-connectivity-toolbox.net/>) to calculate mean strength (the same measure is referred to as mean degree in Chowdhury et al. (2014)^13^), the variance of the strength distribution of all network nodes, average clustering coefficient and average characteristic path length of each functional network. Since networks were weighted and directed, mean strength is the mean of the sum of in- ($a_{ji}$) and out- ($a_{ij}$) strengths to nodes:

$$k_{i}^{out}=\Sigma_{j\in N}a_{ij}$$

$$k_{i}^{in}=\Sigma_{j\in N}a_{ji}$$

$k_{i}^{total}= k_{i}^{in}+k_{i}^{out}$,

whereby 𝑁 is the set of all nodes.

Average clustering coefficient (CC) here, is average ‘intensity’ (geometric mean) of all triangles associated with each node $(i)$:

$$t_{i}=\frac{1}{2}\Sigma_{j,h\in N}(a_{ij}+a_{ji})(a_{ih}+a_{hi})(a_{jh}+a_{hj})$$

${CC}^{original}=\frac{1}{n}\Sigma_{j\in N}\frac{t_{i}}{\left( k_{i}^{out}+k_{i}^{in} \right)\left( k_{i}^{out}+k_{i}^{in}-1 \right)-2\Sigma_{j\in N}{a_{ij}a}_{ji}}$.

For average characteristic path length (PL) (using Floyd-Warshall algorithm), the distance between nodes is the inverse connection strength (1/connection strength) and the average path length is the average shortest path length in the network (whereby $g_{ij}$ is the directed shortest path from $i$ to $j$ and 𝑛 is the number of nodes):

$$d_{ij}=\Sigma_{a_{ij}\in g_{ij}}a_{ij}$$

${PL}^{original}=\frac{1}{n}\Sigma_{j\in N}\frac{\Sigma_{j\in N,j\neq i}d_{ij}}{n-1}$.

Because CC and PL are both sensitive to mean degree and strength, these measures were normalized to the mean CC and PL of 500 random surrogate networks with the same distribution of edge weights:

${CC}^{norm}=\frac{{CC}^{original}}{{CC}^{surrogate}}$,

${PL}^{norm}=\frac{{PL}^{original}}{{PL}^{surrogate}}$.

The small-world index of each network was also calculated according to Humphries and Gurney ^14^:

$S=\frac{{CC}^{norm}}{{PL}^{norm}}$.

##### **Brain Network Ictogenicity (BNI)**

###### **Mathematical model**

A theta model was implemented at each network node to model seizure dynamics.^8,15-17^ The brain activity at node $i$ was represented by a phase oscillator $\theta_{i}$. ‘Resting-state’ is defined as a phase close to a fixed stable phase $\theta^{(s)}$ and an ‘oscillatory state’ as a rotating phase. The resting-state represents normal brain activity, whereas the oscillatory state depicts seizure-like activity. Nodes can transition between these states through a saddle-node on invariant circle (SNIC) bifurcation. The phase oscillator obeys the following ordinary differential equation:

$\dot{\theta}_{i}=(1-\cos\theta_{i})+(1+ \cos\theta_{i})I_{i}(t)$,

where $I_{i}(t)$ is the input current of node $i$ including noisy inputs and the interaction with the other nodes:

$I_{i}\left( t \right)= I_{0}+\xi^{\left( i \right)}\left( t \right)+\frac{K}{N}\sum_{j\neq i} a_{ji}\left[ 1-\cos\left( \theta_{j}-\theta^{(s)} \right) \right]$,

where $I_{0}+\xi^{\left( i \right)}\left( t \right)$is noise, N is the number of nodes, $a_{ji}$ is $j,i^{th}$ entry of the adjacency matrix that encodes the functional network, K is a global scaling factor of the functional network, and $\theta^{(s)}$ is the steady state of the in-neighbor $j$. The noisy inputs ($I_{0}+\xi^{\left( i \right)}\left( t \right)$) represent signals from other areas of the brain outside of the functional network under consideration, which is assumed to follow a Gaussian distribution (with mean $I_{0}=-1.2$ and standard deviation $\sigma^{2}=0.6$, according to previous work^4,16^). Because this study is interested in the influence of functional networks on seizure activity, the chosen parameters ensure that nodes are typically in the resting-state and the transition to seizure-like activity is a result of the network interactions. Lopes et al. (2017)^16^ showed that different choices of $I_{0}$ and $\sigma$ would not quantitatively change the results. The remaining free value, K, will be discussed below.

###### **BNI measure**

BNI^13,15-17^ aims to quantify the propensity of a network to generate seizure-like dynamics in silico. It quantifies the model-generated dynamics with the average fraction of time each node spends in the oscillatory/seizure state:

${BNI}^{*}=\frac{1}{N}\sum_{i} \frac{t_{sz}^{(i)}}{T}$,

where $t_{sz}^{\left( i \right)}$ is the time that node $i$ spent in the oscillatory state during a total simulation time $T$. We used $T$ = 4x10^6^ time steps and the oscillatory state was defined as any activity larger than a threshold as described in Lopes et al. (2017).^16^ The time spent in seizure state ($t_{sz}^{\left( i \right)}$) depends on the global connectivity strength, K, and therefore so does the BNI.^15,16^ To avoid an arbitrary choice of K, we followed the approach described in Lopes et al. (2018)^18^ where we redefined BNI as:

$BNI= \int_{K_{min}}^{K_{max}} {BNI}^{*}(K)dK$,

where $K_{min}$ corresponds to the minimum K value for which $BNI^{*}=0$ across all computed networks and similarly $K_{max}$to the maximum K value that $BNI^{*}=1$. The [$K_{min}, K_{max}$] interval contained 40 equally distributed global scaling (K) values. For each of these values we computed BNI and therefore obtained a distribution of $BNI^{*}$ values that results in a curve for each EEG (Figure S 1). This approach allows for comparison between individuals and avoids arbitrary choice of K. To quantify the variation of $BNI^{*}$ between $K_{min}$ and $K_{max}$, the area under the BNI curve generated for each EEG epoch was calculated using the MATLAB ‘trapz’ function which performs numerical integration via the trapezoidal method.

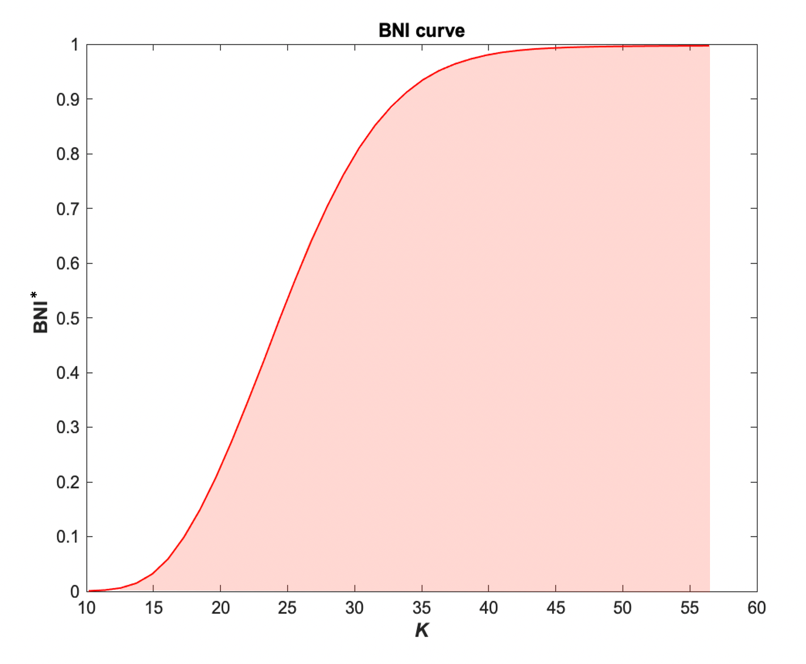

**Figure S 1** - Example Brain Network Ictogenicity (BNI) curve generated by calculating the BNI of a single functional network.

#### **Supplementary Figures & Tables**

##### **Influence of ICA on EEG frequency measures**

ICA was used to remove artefacts in 39/95 (41%) of control EEGs and 86/194 (44%) of JME EEGs. There was extremely high correlation between relative PSD in each frequency band in epochs before and after they underwent artifact removal ICA (Figure S 2 & Table S 1). Therefore, I used the epochs which had undergone ICA for the following frequency analysis.

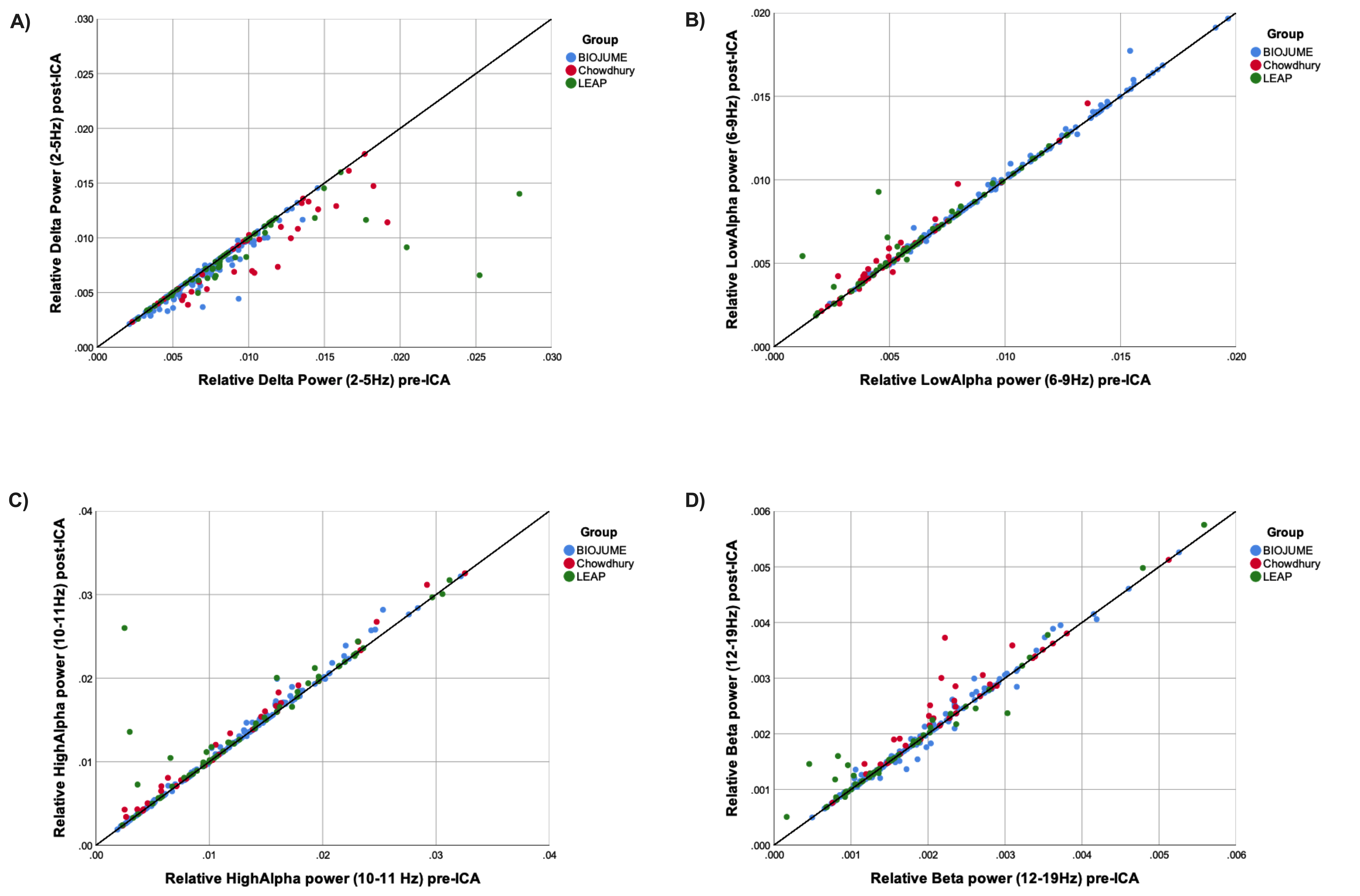

**Figure S 2** - Relative power spectral density measures in **A**) Delta, **B**) Low-alpha **C**) High-alpha and **D**) Beta, pre- and post-independent component analysis (ICA).

**Table S 1 -** Pearson's correlation coefficients for pre- and post-ICA relative power spectral density in each frequency band, for each EEG group.

| **EEG Group** | **Delta (2-5Hz)** | **Low-alpha (6-9Hz)** | **High-alpha (10-11Hz)** | **Beta (12-19Hz)** |
| --- | --- | --- | --- | --- |
| **BIOJUME** | 0.97 | 1.0 | 1.0 | 0.99 |
| **LEAP** | 0.73 | 0.95 | 0.89 | 0.98 |
| **Chowdhury** | 0.93 | 0.98 | 1.0 | 0.95 |

##### **BIOJUME site variation in clustering coefficient**

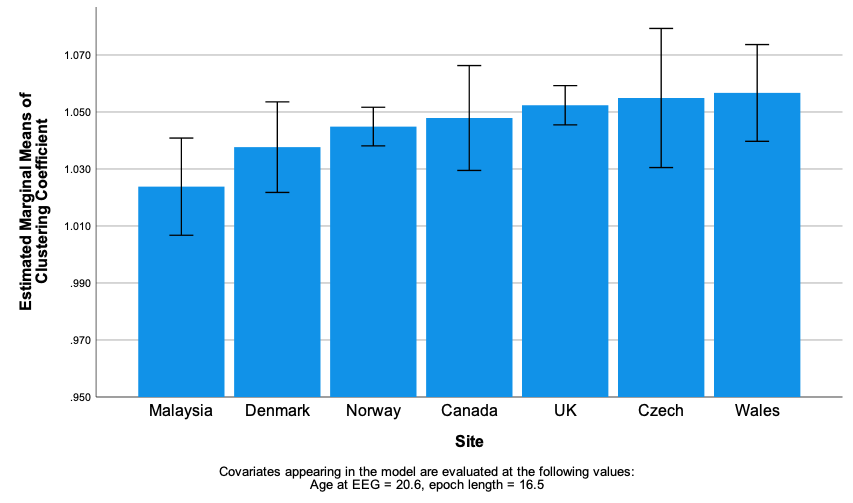

**Figure S 3** - Estimated marginal means of clustering coefficient in JME EEGs stratified by site, controlling for age at EEG and epoch length. Error bars show 95% confidence intervals.

##### **Power spectral density in JME vs Controls**

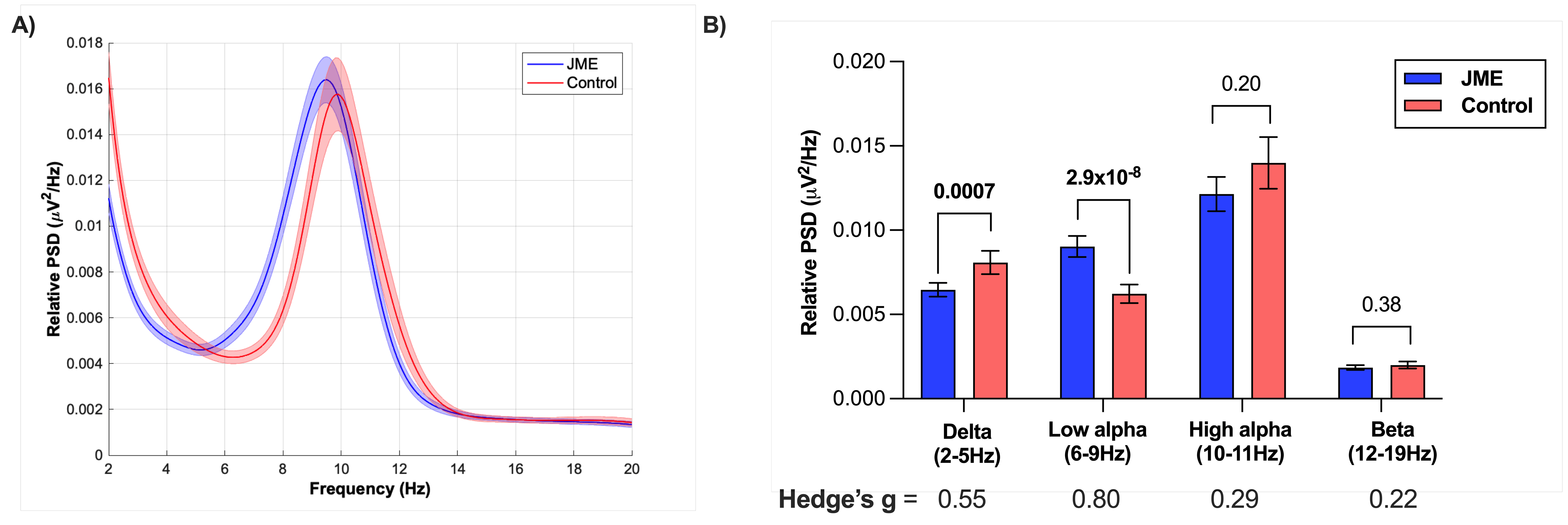

**Figure S 4 - A)** Relative power spectral density (PSD) plots of JME and control EEGs between 2-20Hz. Solid line is the subgroup mean and shaded area represents 95% confidence intervals. **B)** Quantification of relative PSD in EEG frequency bands between control and JME EEGs. Values above bars represent p-values from Mann-Whitney U test, corrected for the 4 frequency bands using Bonferroni-holm. Hedge’s g values for each comparison are below bars.

##### **Regression models**

**Table S 2** - Multiple linear regression model results for **A)** relative delta power spectral density (PSD), **B)** relative low-alpha PSD, **C)** relative high-alpha PSD and **D)** relative beta PSD. Bold p-values = <0.05. For binary variables categories are presented as they were coded (0/1).

| **A) Delta** | | | | |  | **B) Low-alpha** | | | | |
| --- | --- | --- | --- | --- | --- | --- | --- | --- | --- | --- |
| **Variables** | **Unstandardized coefficient** | | **T** | **P-value** |  | **Variables** | **Unstandardized coefficient** | | **T** | **P-value** |
|  | B | Std Error |  |  |  |  | B | Std Error |  |  |
| **Control/JME** | -0.0023 | 0.00039 | -6.04 | **6.0x10^-9^** |  | **Control/**  **Untreated JME** | 0.0033 | 0.000616 | 5.29 | **2.9x10^-7^** |
| **Age** | -0.0002 | 2.1x10^-5^ | -8.13 | **5.0x10^-6^** |  | **Control/**  **Treated JME** | 0.0022 | 0.000556 | 3.99 | **9.1x10^-5^** |
| **Epoch length** | -0.00014 | 5.1x10^-5^ | -2.34 | **0.0076** |  | **Age** | -1.6x10^-5^ | 0.000028 | -0.99 | 0.57 |
| N =236, adjusted r2 =0.19 | | | | |  | **Epoch length** | -2.3x10^-5^ | 0.000067 | -0.34 | 0.73 |
|  |  |  |  |  |  | N =225, adjusted r^2^ =0.15 | | | | |
| **C) High-alpha** | | | | |  | **D) Beta** | | | | |
| **Variables** | **Unstandardized coefficient** | | **T** | **P-value** |  | **Variables** | **Unstandardized coefficient** | | **T** | **P-value** |
|  | B | Std Error |  |  |  |  | B | Std Error |  |  |
| **Age** | 8.3x10^-5^ | 5.3x10^-5^ | 1.56 | 0.12 |  | **Age** | 2.4x10^-5^ | 8.0x10^-5^ | 3.09 | **0.0023** |
| **Epoch length** | 0.00017 | 0.00013 | 1.32 | 0.19 |  | **JME/Chowdhury** | 0.00038 | 0.00018 | 2.06 | **0.041** |
| **Control/JME** | -0.00094 | 0.00097 | -0.97 | 0.33 |  | **JME/LEAP** | -9.0x10^-5^ | 0.00014 | -0.63 | 0.53 |
| N =236, adjusted r^2^ =0.02 | | | | |  | **Epoch length** | -4.9x10^-6^ | 1.7x10^-5^ | -0.29 | 0.77 |
|  |  |  |  |  |  | N =236, adjusted r^2^ =0.09 | | | | |

**Table S 3** - Results of a multiple linear regression model of log_10_ alpha shift. Bold p-values = <0.05. Control/JME coded as 0/1*.*

**Log_10_ Alpha Shift**

| Variables | Unstandardized coefficient. | | T | P value |
| --- | --- | --- | --- | --- |
|  | B | St Error |  |  |
| Control/JME | 1.28 | 0.48 | 2.66 | **0.009** |
| EEG time* Control/JME | -2.4x10^-8^ | 9.7x10^-9^ | -2.5 | **0.013** |
| Age | -0.006 | 0.005 | -1.38 | 0.17 |
| EEG time | 1.1x10^-8^ | 8.0x10^-9^ | 1.35 | 0.18 |
| Mean epoch length | 0.0002 | 0.0083 | 0.03 | 0.98 |
| N =172, adjusted r^2^ =0.05 | | | | |

**Table S 4 -** Multiple linear regression model of peak alpha frequency. Bold p-values= <0.05. Control/JME coded as 0/1.

**Peak alpha frequency**

| Variables | Unstandardized coefficient. | | T | P value |
| --- | --- | --- | --- | --- |
|  | B | St Error |  |  |
| Control/JME | -3.18 | 1.00 | -3.18 | **0.0018** |
| EEG time* Control/JME | 5.8x10^-5^ | 2.0x10-5 | 2.86 | **0.0048** |
| EEG time | -3.7x10^-5^ | 1.7x10-5 | -2.24 | **0.026** |
| Sex | 0.24 | 0.14 | 1.77 | 0.078 |
| Age | 0.0012 | 0.0095 | 0.12 | 0.90 |
| Epoch length | 0.005 | 0.018 | -0.03 | 0.98 |
| N =168, adjusted r^2^ =0.06 | | | | |

**Table S 5** - Results of multiple linear regression analysis of **A)** mean strength, **B)** Mean strength variance, **C)** clustering coefficient, **D)** Path length and **E)** Small-world index of functional networks derived from EEGs. For binary variables categories are presented as they were coded (0/1).

| **A) Mean strength** | | | | |  | **B) Mean strength variance** | | | | |
| --- | --- | --- | --- | --- | --- | --- | --- | --- | --- | --- |
| Variables | Unstandardized coefficient. | | T | P value |  | Variables | Unstandardized coefficient. | | T | P value |
|  | B | St Error |  |  |  |  | B | St Error |  |  |
| Control/JME | 0.832 | 0.17 | 4.89 | **1.9x10^-6^** |  | JME/Chowdhury | -1.15 | 0.26 | -4.31 | **2.4x10^-5^** |
| Epoch length | 0.068 | 0.022 | 2.96 | **0.003** |  | Age | 0.02 | 0.012 | 2.25 | **0.025** |
| Age | 0.027 | 0.009 | 2.84 | **0.005** |  | Epoch length | -0.02 | 0.023 | -0.89 | 0.38 |
| N =235, adjusted r^2^ =0.12 | | | | |  | JME/LEAP | 0.16 | 0.2 | 0.82 | 0.41 |
|  |  |  |  |  |  | N =229, adjusted r^2^ =0.08 | | | | |
| **C) Clustering coefficient** | | | | |  | **D) Path Length** | | | | |
| Variables | Unstandardized coefficient. | | T | P value |  | Variables | Unstandardized coefficient. | | T | P value |
|  | B | St Error |  |  |  |  | B | St Error |  |  |
| Epoch length | -0.004 | 0.0005 | -8.6 | **1.3x10^-15^** |  | JME/Chowdhury | -0.061 | 0.0065 | -2.43 | **0.016** |
| JME/LEAP | 0.021 | 0.004 | 4.97 | **1.3x10^-6^** |  | Epoch length | 0.00039 | 0.00059 | 0.66 | 0.51 |
| JME/Chowdhury | -0.0064 | 0.005 | -1.21 | 0.23 |  | Age | 0.00018 | 0.00027 | 0.66 | 0.51 |
| Age | -0.0001 | 0.0002 | -0.47 | 0.64 |  | JME/LEAP | 0.0013 | 0.0051 | 0.25 | 0.80 |
| N = 233, adjusted r^2^ =0.33 | | | | |  | N =231, adjusted r^2^ =0.01 | | | | |
| **E) Small-world index** | | | | |  |  |  |  |  |  |
| Variables | Unstandardized coefficient. | | T | P value |  |  |  |  |  |  |
|  | B | St Error |  |  |  |  |  |  |  |  |
| Epoch length | -0.0046 | 0.00056 | -8.14 | **2.7x10^-14^** |  |  |  |  |  |  |
| JME/LEAP | 0.017 | 0.0049 | 3.57 | **0.0004** |  |  |  |  |  |  |
| JME/Chowdhury | 0.0098 | 0.0061 | 1.61 | 0.12 |  |  |  |  |  |  |
| Age | -0.00033 | 0.00026 | -1.28 | 0.20 |  |  |  |  |  |  |
| N =228, adjusted r^2^ =0.25 | | | | |  |  |  |  |  |  |

**Table S 6** - Multiple linear regression model of Brain Network Ictogenicity (BNI) area under the curve (AUC). Control/JME coded as 0/1.

**BNI AUC**

| Variables | Unstandardized coefficient. | | T | P value |
| --- | --- | --- | --- | --- |
|  | B | St Error |  |  |
| Control/JME | 2.96 | 0.57 | 5.17 | **5.1x10^-7^** |
| Age | 0.11 | 0.03 | 3.60 | **0.0004** |
| Mean epoch length | 0.16 | 0.07 | 2.11 | **0.036** |
| N =235, adjusted r^2^ =0.13 | | | | |

##### **Biomarker cut off point – low-alpha PSD**

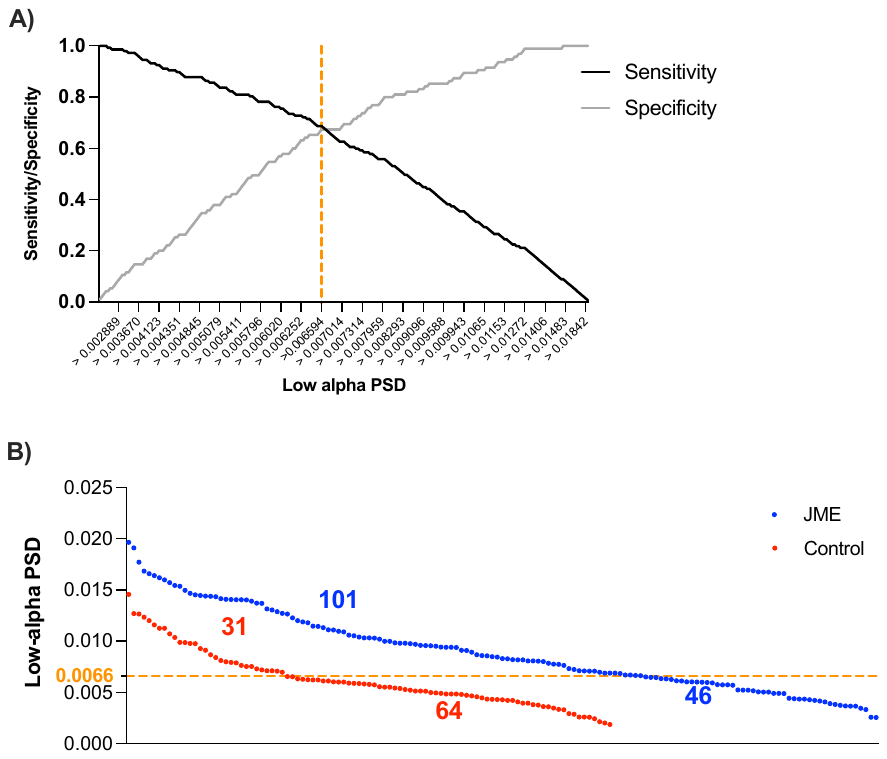

**Figure S 5 -** **A)** A graphical representation of our cut-off choice for pathogenic low-alpha power spectral density (PSD). We chose the cut-off value with the highest sensitivity (0.69) and specificity (0.67). This value is marked by the orange line at 0.0066. **B)** The distribution of low-alpha PSD in JME (blue) and controls (red) and the numbers within the two groups above and below the proposed threshold. Each circle represents an individual. Based on these numbers the Positive Predictive Value=77% and the Negative Predictive Value=58%.

### **Appendix**

Sites and site investigators in the BIOJUME consortium.

| **Country** | **Site** | **Site principal investigators (PIs) and research staff** |
| --- | --- | --- |
| Canada | SickKids Hospital, Toronto | Lisa Strug (PI), Naim Panjwani, Fan Lin |
|  | Toronto Western Hospital | Danielle Andrade (PI) |
| Czech Republic | Charles University | Jana Zarubova (PI), Zuzana Šobíšková |
| Denmark | Danish National Epilepsy Centre | Guido Rubboli (PI), Rikke S. Møller, Elena Gardella |
|  | Syddansk Universitet | Christoph P. Beier (PI), Joanna Gesche |
| Estonia | Tallinn Children's Hospital | Inga Talvik (PI) |
| Italy | Commissione Genetica | Pasquale Striano (PI), Alessandro Orsini |
|  | University of Catania | Andrea Pratico (PI) |
| Malaysia | University of Malaya | Choong Yi Fong (PI), Ching Ching Ng, Kheng Seang Lim |
| Norway | Vestre Viken Health Trust | Jeanette Koht, Kaja K. Selmer, Marte Syvertsen (Co-PIs) |
| UK | Airedale NHS Foundation Trust | Pronab Bala (PI), Amy Kitching |
|  | Ashford and St. Peter’s Hospitals NHS Foundation Trust | Kate Irwin (PI), Lorna Walding, Lynsey Adams |
|  | Bradford Teaching Hospitals NHS Foundation Trust | Uma Jegathasan (PI), Rachel Swingler, Rachel Wane |
|  | Brighton and Sussex University Hospitals NHS Trust | Julia Aram (Co-PI), Nikil Sudarsan (Co-PI), Dee Mullan, Rebecca Ramsay, Vivien Richmond, Mark Sargent, Paul Frattaroli |
|  | Calderdale and Huddersfield Foundation Trust | Matthew Taylor (PI), Marie Home, Sal Uka, Susan Kilroy, Tonicha Nortcliffe, Halima Salim, Kelly Holroyd |
|  | Cardiff & Vale University Health Board | Khalid Hamandi (PI), Alison McQueen, Dympna Mcaleer |
|  | County Durham and Darlington NHS Foundation Trust | Dina Jayachandran (PI), Dawn Egginton, |
|  | Croydon Health Services NHS Trust | Bridget MacDonald (PI), Michael Chang |
|  | Cwm Taf Morgannwg University Health Board | David Deekollu (Co-PI), Alok Gaurav (Co-PI), Caroline Hamilton, Jaya Natarajan |
|  | Dartford and Gravesham NHS Trust | Shane Delamont (PI), Carmel Stuart, Imogen Hayes |
|  | East and North Hertfordshire NHS Trust | Inyan Takon (PI), Janet Cotta |
|  | East Kent Hospitals University NHS Foundation Trust | Nick Moran (PI), Jeremy Bland |
|  | East Lancashire Hospitals NHS Trust | Rosemary Belderbos (PI), Heather Collier, Joanne Henry, Matthew Milner, Sam White |
|  | Guy's and St Thomas' NHS Foundation Trust | Michalis Koutroumanidis (PI), Javier Peña Ceballos, William Stern |
|  | King's College Hospital NHS Foundation Trust | Mark P. Richardson (Co-PI), Jennifer Quirk (Co-PI), Javier Peña Ceballos, Anastasia Papathanasiou |
|  | King's College London | Deb K. Pal (PI), Mark P. Richardson, Holly Crudgington, Anna Hall, Amber Collingwood, Amy Shakeshaft, Ioannis Stavropoulos, Anna Smith, Robert McDowall, Sophie Bayley |
|  | Kingston Hospital NHS Foundation Trust | Dora Lozsadi (PI), Andrew Swain, Charlotte Quamina, Jennifer Crooks |
|  | Lancashire Teaching Hospitals NHS Foundation Trust | Tahir Majeed (PI), Sonia Raj, Shakeelah Patel, Michael Young |
|  | Leeds Teaching Hospitals NHS Trust | Melissa Maguire (Co-PI), Munni Ray (Co-PI), Caroline Peacey, Linetty Makawa, Asyah Chhibda, Eve Sacre, Shanaz Begum |
|  | Manchester University NHS Foundation Trust | Lap Yeung (Co-PI), Claire Holliday, Louise Woodhead, Karen Rhodes |
|  | Newcastle upon Tyne Hospitals NHS Foundation Trust | Rhys Thomas (Co-PI), Shan Ellawela (Co-PI), Joanne Glenton, Verity Calder, John Davis, Paul McAlinden, Sarah Francis |
|  | NHS Grampian | Karen Lanyon (Co-PI), Graham Mackay (Co-PI), Elma Stephen (Co-PI), Coleen Thow, Margaret Connon |
|  | NHS Tayside | Martin Kirkpatrick (PI), Susan MacFarlane, Anne Macleod, Debbie Rice |
|  | North Tees and Hartlepool NHS Foundation Trust | Siva Kumar (PI), Carolyn Campbell, Vicky Collins |
|  | Nottingham University Hospitals NHS Trust | William Whitehouse (PI), Christina Giavasi (PI), Boyanka Petrova, Thomas Brown, Catie Picton, Michael O'Donoghue, Charlotte West, Helen Navarra |
|  | Portsmouth Hospitals NHS Trust | Seán J. Slaght (PI), Catherine Edwards, Andrew Gribbin, Liz Nelson, Stephen Warriner |
|  | Royal Free London NHS Foundation Trust | Heather Angus-Leppan (PI), Loveth Ehiorobo, Bintou Camara, Tinashe Samakomva |
|  | Salford Royal NHS Foundation Trust | Rajiv Mohanraj (PI), Vicky Parker |
|  | Sandwell & West Birmingham Hospitals NHS Trust | Rajesh Pandey (PI), Lisa Charles, Catherine Cotter |
|  | Sheffield Children's NHS Foundation Trust | Archana Desurkar (PI), Alison Hyde, Rachel Harrison |
|  | Sheffield Teaching Hospitals NHS Foundation Trust | Markus Reuber (PI), Rosie Clegg, Jo Sidebottom, Mayeth Recto, Patrick Easton, Charlotte Waite, Alice Howell, Jacqueline Smith, Rosie Clegg |
|  | Southport and Ormskirk Hospital NHS Trust | Shyam Mariguddi (PI), Zena Haslam |
|  | St George's University Hospitals NHS Foundation Trust | Elizabeth Galizia (PI), Hannah Cock, Mark Mencias, Samantha Truscott, Deirdre Daly, Hilda Mhandu, Nooria Said |
|  | Swansea University Medical School and Swansea Bay University Healthboard | Mark Rees (PI), Seo-Kyung Chung, Owen Pickrell, Beata Fonferko-Shadrach, Mark Baker |
|  | Taunton & Somerset NHS Foundation Trust | Amy Whiting (PI), Kirsty O’Brien |
|  | The Mid Yorkshire Hospitals NHS Trust | Fraser Scott (Co-PI), Naveed Ghaus (Co-PI), Gail Castle, Jacqui Bartholomew, Ann Needle, Julie Ball, Andrea Clough |
|  | The Royal Wolverhampton NHS Trust | Shashikiran Sastry (PI), Charlotte Busby |
|  | The Walton Centre NHS Foundation Trust | Amit Agrawal (PI), Debbie Dickerson, Almu Duran |
|  | University Hospitals Birmingham NHS Foundation Trust | Muhammad Khan (PI), Laura Thrasyvoulou, Eve Irvine, Sarah Tittensor, Jacqueline Daglish |
|  | University Hospitals of Derby and Burton NHS Foundation Trust | Sumant Kumar (PI), Claire Backhouse, Claire Mewies |
|  | University Hospitals Plymouth NHS Trust | Rahul Bharat (PI), Sarah-Jane Sharman |
|  | Walsall Healthcare NHS Trust | Darwin Pauldhas (PI), Sharon Kempson, Lisa Richardson, Lynn Hawkins |
|  | West Suffolk NHS Foundation Trust | Arun Saraswatula (PI), Helen Cockerill |
| USA | Nationwide Children's Hospital, Ohio | David A. Greenberg (PI) |
